# Aesthetic evaluation of the need for orthodontic treatment – Perception among university students

**DOI:** 10.1101/2020.12.22.20244806

**Authors:** Luís Alves, Anca Mesaros, Maria João Ponces, Maria Cristina Figueiredo Pollmann

## Abstract

**Introduction:** Aesthetics is a relevant part of procedures in healthcare, often influencing treatment planning in tandem with a healthy function. Orthodontic treatment (OT) is one of many solutions and is, sometimes, wanted purely for aesthetic reasons. In 1989, Brook and Shaw proposed the Index of Orthodontic Treatment Need (IOTN), which has been largely used.

This study aims to verify the main motivations of university students to look for OT and, based on the Aesthetic Component of the IOTN, weigh the aesthetics influence to seek for it. It was compared the opinion of students from various areas of study - Dentistry, Science and Nature (ScN), Arts and Humanity (AtH) - at the beginning (Initiated Students) and end (Advanced Students) of their graduation; the same question was also analysed taking into account their nationalities and training schools.

**Materials and Methods:** In a collaboration between the University of Porto (Portugal) and the University of Medicine and Pharmacy of Cluj-Napoca (Romania), a sample of 1071 individuals was gathered. Participants responded to an online survey, based on IOTN pictures, about what would motivate them to seek OT. The ratings were analysed using the T-Test and alpha error of 5%.

**Results:** The results showed that the total Dentistry students registered a higher Oral Esthetical Sensibility (OES) than the total ScN and AtH students. All groups, except Dentistry Advanced Students, registered a higher OES for Self-Perception than for Perception of Others.

**Discussion:** Among other factors, the pictures used from the IOTN, taken in 1989, may have influenced the participants’ responses. However, it is the most used index, and it is validated.

**Conclusion:** In conclusion, for the studied populations, the main motivations for OT demand are primarily and respectively in this order: functional reasons, doctor’s advice, and aesthetical reasons. OES is influenced by Dentistry studies, specifically in Advanced Students. OES is not influenced by the country of the students’ origin nor the country they are graduating at.

## Introduction

### Cultural perception of Aesthetics

Aesthetic standards are subjective and influenced by many factors, such as fashion, cultural traditions, personalities, self-esteem, peers, or, even, ideologies.

On human physical beauty concepts and practices aiming to achieve it, the objectives are set by the standards of a group of people rather than by science. On the other hand, when one researches the definition of “health”, in a holistic meaning, it is a concept that nowadays is defined by the World Health Organization (WHO) as “a state of complete physical, mental and social well-being and not merely the absence of disease or infirmity”. This means that although Aesthetics effectively plays a role, the concept of human physical beauty will need to be defined according to the individual’s global general health priorities. Currently, good health practices are typically based on scientific methods. Conciliating the procedures targeted at human physical aesthetics with the best individual’s state of general health has become a challenge. Among other factors, the evolution of knowledge and the increase in economic power, motivated a large demand for orthodontic treatment (OT), in a desire for better functional aesthetics. Noticing that someone is using a fixed system or a post-treatment retention to prevent the receding of the OT is not uncommon in our daily lives. ^(1-8)^

Moreover, we commune in a paradigm undeniably marked by social media and the Internet. Fulfilling one’s wishes in a rapid manner is increasingly, being taken as the rule and not the exception. Ideas and decision making will typically be influenced by the opinion of a third party who used or tested a specific beauty product or solution. On that basis, subjective pressure on aesthetics is a very relevant component of determining the “necessity” of orthodontic treatment and, which health professionals cannot be alienated from, nor its dimension and impact. ^(1, 3, 7-14)^

### The Index of Orthodontic Treatment Need

Plenty of indexes have been formerly proposed to the scientific community to facilitate health practitioners in understanding the need for orthodontic treatment in different cases of malocclusions. The Index of Orthodontic Treatment Need (IOTN), published in 1989 by Brook and Shaw and massively used by the British National Health System (NHS) includes two distinct components: the Dental Health Component (DHC) and the Aesthetic Component (AC). These correlate the need for orthodontic treatment with different characteristics regarding oral health, such as occlusion, function, and oral aesthetics. The index was created with the purpose of establishing a coherent prioritisation for orthodontic treatment needs, that is, the bigger the need for treatment, the higher priority.

If a patient is considered for treatment in the DHC grading, the orthodontist will then look at the appearance of the smile and give a second score, according to the Aesthetic Component, so the treatment is not for purely aesthetic reasons. The AC has a scale of 10 pictures, arranged by non-dental related people, showing different oral cavities of different levels of attractiveness. ^(5, 15, 16)^

### The Perception of University Students

In what regards the need for orthodontic treatment, the correlation between self-perception and the professional’s judgement may differ. Similarly, it may differ among Health students in the beginning and at the end of their graduation. This is a gradual process, especially in the case of Dentistry students who already start their course with a formed opinion about what is beautiful and sound. Later, the students acquire a bigger competence and aptitude that typically leads towards moderate opinions, as opposed to their previous stronger ones, and, with time, the students might even develop new and renovated convictions. Sometimes, they even start defending the perceptions that were less popular to them before in the beginning. Reportedly, students in the first year of Dentistry might give higher importance to aesthetics, when compared to final students. Since the paradigm of Science is always mutating, a few questions are raised: Is this a fact or given knowledge? Does it depend on their previous education? Is this a consequence of the learning in their respective faculty? Is their cultural background as determinant as it may seem? ^(17)^

### Objectives

Based on the Aesthetic Component of the IOTN, this study aims to find out what might influence those perceptions and how it takes place, in a population of University Students. For this purpose, the project was divided into two parts:

In the first part, the aim was to determine if the Portuguese and Romanian cultures have a bearing on it; as well as if the learning of Dentistry affects it. Also, if the perceptions of International Dentistry Students of UMF are influenced by their own culture or by university culture where they are graduating.

In the second part, the goal was to ascertain how the Academic Degree and the Area of Studies may have a role on Student’s perceptions, specifically those in the fields of Science and Nature (allegedly more objective thinking) and Art and Humanity (allegedly more critical aesthetic thinking), as well as Dentistry.

The hypotheses to be studied are: Is students’ perception influenced by the country they grew up or the country they are studying? Is it influenced by their area of studies or their academic maturity? Is students’ Self-Perception different than their Perception of Others?

## Material and Methods

### Introduction

The following research, a collaboration between the University of Porto (Portugal) and the University of Medicine and Pharmacy (UMF) “Iuliu Ha□ieganu” of Cluj-Napoca (Romania), takes part of a study about University Students’ opinion on Orthodontic Treatment. Students were invited to answer, voluntarily, to a series of questions (Annex 1). The study included students from all the 14 Faculties and Schools of UP and the Romanian Universities of Cluj-Napoca UMF, UTC-N, UBB, UAD, and USAMV Cluj-Napoca.

We used the CONSORT checklist when writing our report.^(18)^

### Literature revision

The research and the premises that gave rise to this investigation were obtained from the Database PubMed with the following keywords: “Beauty”, “Esthetics/psychology”, “Quality of Life/psychology”, “IOTN”, “Aesthetic Component of IOTN”, “orthodontics AND economy”, “(orthodontic treatment) AND self-perception (orthodontic treatment) AND self-perception”, “(orthodontic treatment) AND social media”, “(orthodontic treatment) AND quality of life”, (“Respiration”[Mesh]) AND “Orthodontic Appliances”[Mesh], (orthodontics) AND (pediatrics).

The search was trunked to articles published from 2008 to 2020, with the exception for the Aesthetic Component of Index of Orthodontic Treatment Need, article written in 1989.

### Ethical Considerations

The inquiry was completely anonymous, no name or identification was asked from the participants. No one was submitted to any kind of risk or harm nor any kind of payment or gratification was granted to those who answered.

The pictures used in the Clinical Cases were taken from the 1989 IOTN, by Brook and Shaw.

The inquiry was approved by the Ethics Commission of FMDUP and the UP’s Data Protection Office, being the verdict valid to the Dean’s office of UMF (annexes section).

### Sample

The inclusion criterion for the general sample was to be a student, born between the years of 1980 and 2002, and enrolled in any of the faculties and schools of the University of Porto (Portugal) or any of the Cluj-Napoca’s Universities (Romania).

The general sample of university students was then divided taking into consideration two main groups with the following inclusion criteria:

- Initial Studies Group (IS): all students from the first to the penultimate year of their course, were considered;
- Advanced Studies Group (AS): all graduating students or at least in the 5^th^ year and all those attending postgraduate or doctoral studies were considered. Each of these groups was sub-divided into
- Science and Nature student
- Art and Humanity student
- Dentistry student

### Methodology

The participants were invited to respond an inquiry created on Google Forms and shared via email. (Annex Section)

The participants should fill in their opinion, rating from 1 to 5 as “most certainly wouldn’t” and “most certainly would”, respectively, on the looking for OT, in four motivational fields: Doctor’s Advice, Functional Reason, Aesthetic Reason and Fashion.

The same rating system was applied to the following motivational perspectives: “If this was your mouth, would you seek Orthodontic Treatment?” and “If this was someone’s mouth, who asked for your opinion, would you recommend Orthodontic Treatment?”.

The Clinical Cases were taken from the Aesthetic Component of the IOTN as seen in Image 1.

**Image 1.**
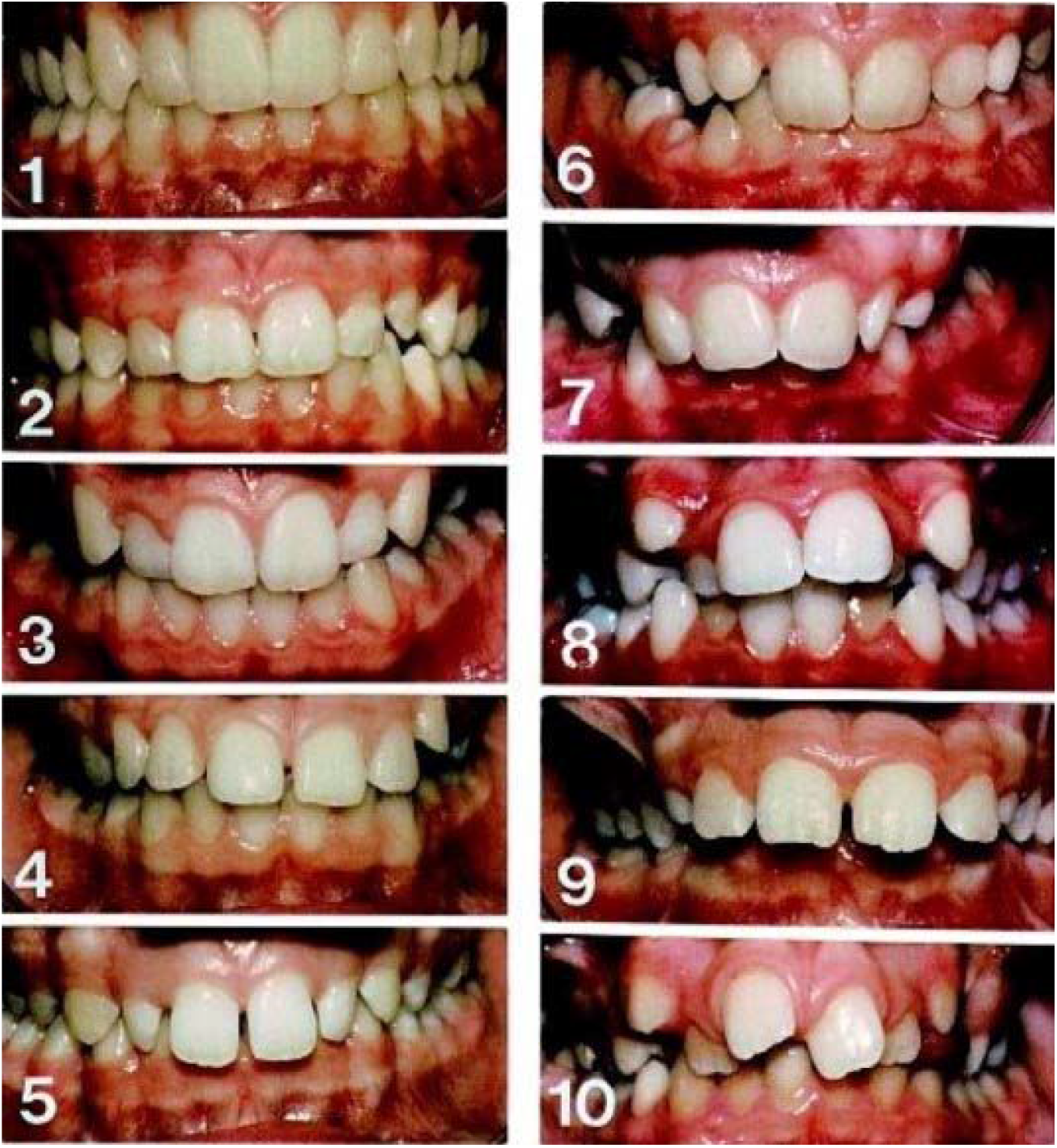
Aesthetic Component of the Index of Orthodontic Treatment Need; Source: Brook PH, Shaw WC. The development of an index of orthodontic treatment priority. Eur J Orthod. 1989;11(3):309-20; Adapted.

The survey took place between dates 15^th^ of January and 15^th^ April. The data was collected and recorded as a Microsoft Office Excel document.

### Statistical Analysis

The data has been entered into a database supported by *SPSS 26® for Windows*. A descriptive analysis of the Sample was performed focusing on Birth Year, Gender, Course of Studies, Academic Degree and Motivations, i.e., how much does Doctor’s Advice, Aesthetic, Function, and Fashion influence the search for Orthodontic Treatment. The applied test for the comparison was T-Test for Independent Samples and for Paired Samples.

Concerning the answers that the participants gave to the AC for Self-Perception and for Perception of Others, the statistical interpretation was made as follows:

The ratings, from 1 to 5, for each of the 10 pictures of the AC were summed resulting in a single rating (per observer) with a score ranging from a minimum of 10 to a maximum of 50. Thus, was created the Variable of Oral Esthetical Sensibility (OES) measured from 10-50.

Henceforth, used for the Self-Perception and another for the Perception of Others. (Image 2)

**Image 2.**
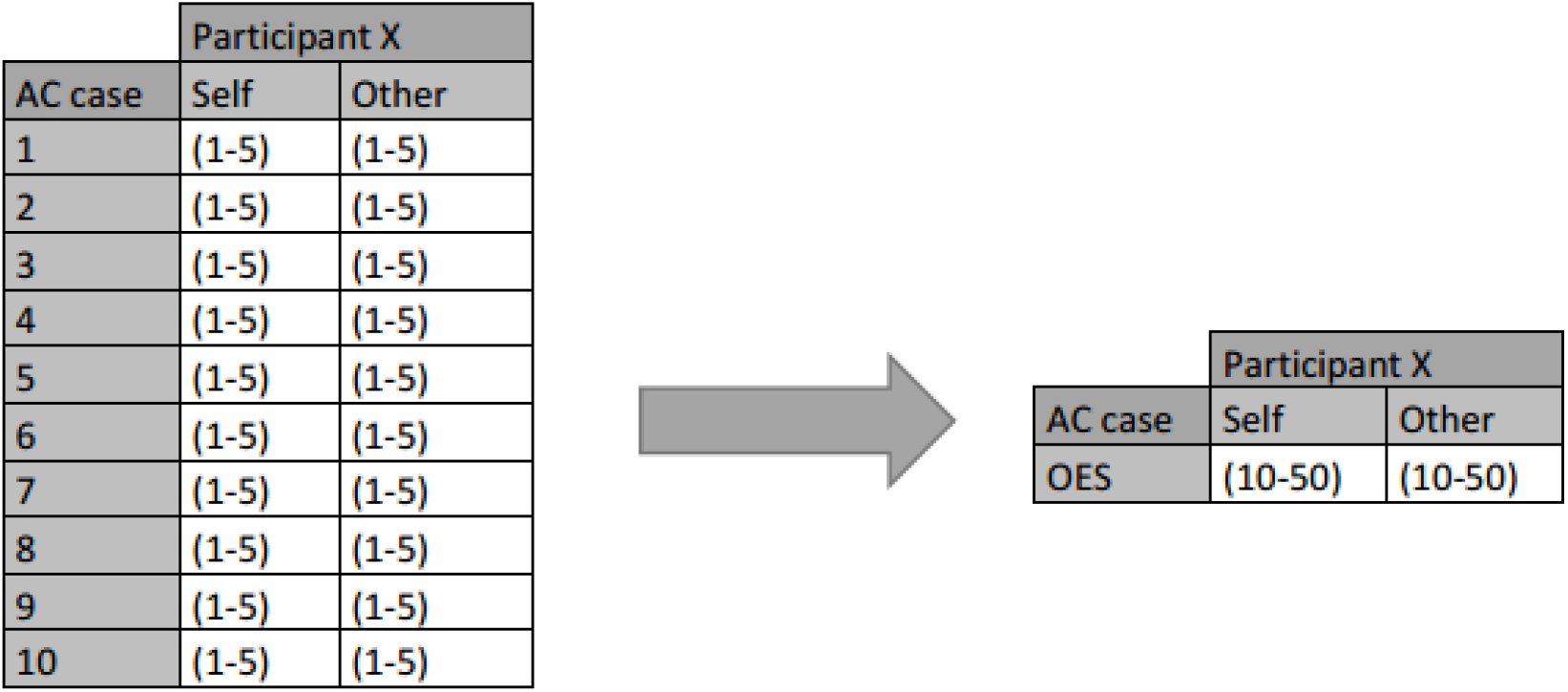
Developing the Oral Esthetical Sensibility

The comparisons between the OES mean values for both the Others Perception and of the Self were for the following groups:

1. Portuguese and Romanian answers
  a. Portuguese and Romanian Dentistry Students
2. Romanian Dentistry Students and International Dentistry Students in UMF
3. All Initiated Students and all Advanced Students
  a. In the generality of University Students
  b. In Dentistry
  c. In Art and Humanity
  d. In Science and Nature
4. Art and Humanity and Science and Nature
5. Dentistry Students and All Other Students
  a. Generality
  b. Initiated Studies
  c. Advanced Studies

## Results

### Sample

The interval of ages of all the n=1071 students varies between 18 and 40 and the mean age is 23. The birth years between 1995 and 2001 compose 81,1% of all participants.

For the Initiated Studies Group, all participants from 1st to 4th years in their respective Course of Studies were included. The group had n=619 (57,8%), the mean age was 21, and the birth year ranging from 1981 to 2002.

In the Advanced Studies Group were included the 5th, last years, and post-graduate students, n=452 (42,2%) in Advanced Studies, mean age was 25, and the birth year ranging from 1980 to 1999.

Grouped by Course of Studies: Dentistry n=186, 40,9% in Initiated Studies; Arts and Humanity n=209, 64,1% in Initiated Studies; Science and Nature n=676, being 60,5% in Initiated Studies.

#### First Part of the Study

For the first part, only 158 respondents were considered, adapted from the original database, to homogenize, the Portuguese and Romanian occurrences, each with n=50. (Annex Section)

In this adapted sample, the mean age of Portuguese students is 23 years, 37 female and 13 male participants whereas the Romanian the mean age is 25, being 41 female and 9 male.

The International Dentistry Students in UMF has n=58, with the mean age of 24, includes 31 female and 27 male students.

#### Second Part of the Study

For the second part, the whole database n=1071 was used.

### Motivations for Orthodontic Treatment

All 1071 participants were asked to choose from 1 to 5 how much doctor’s advice, aesthetic, functional, and fashion reasons influence their drive to search for Orthodontic Treatment. In crescent order, 1 meant “most certainly wouldn’t” and 5 “most certainly would”.

Regarding Doctor’s Advice, 54,9% of the participants rated 5. Aesthetic Reasons present a rate of 5 for 41,1%, Functional Reasons were rated 5 by 61,3% of the participants. For Fashion, 68,8% rated 1, however 12,3% rated 3 or above.

**Image 3.**
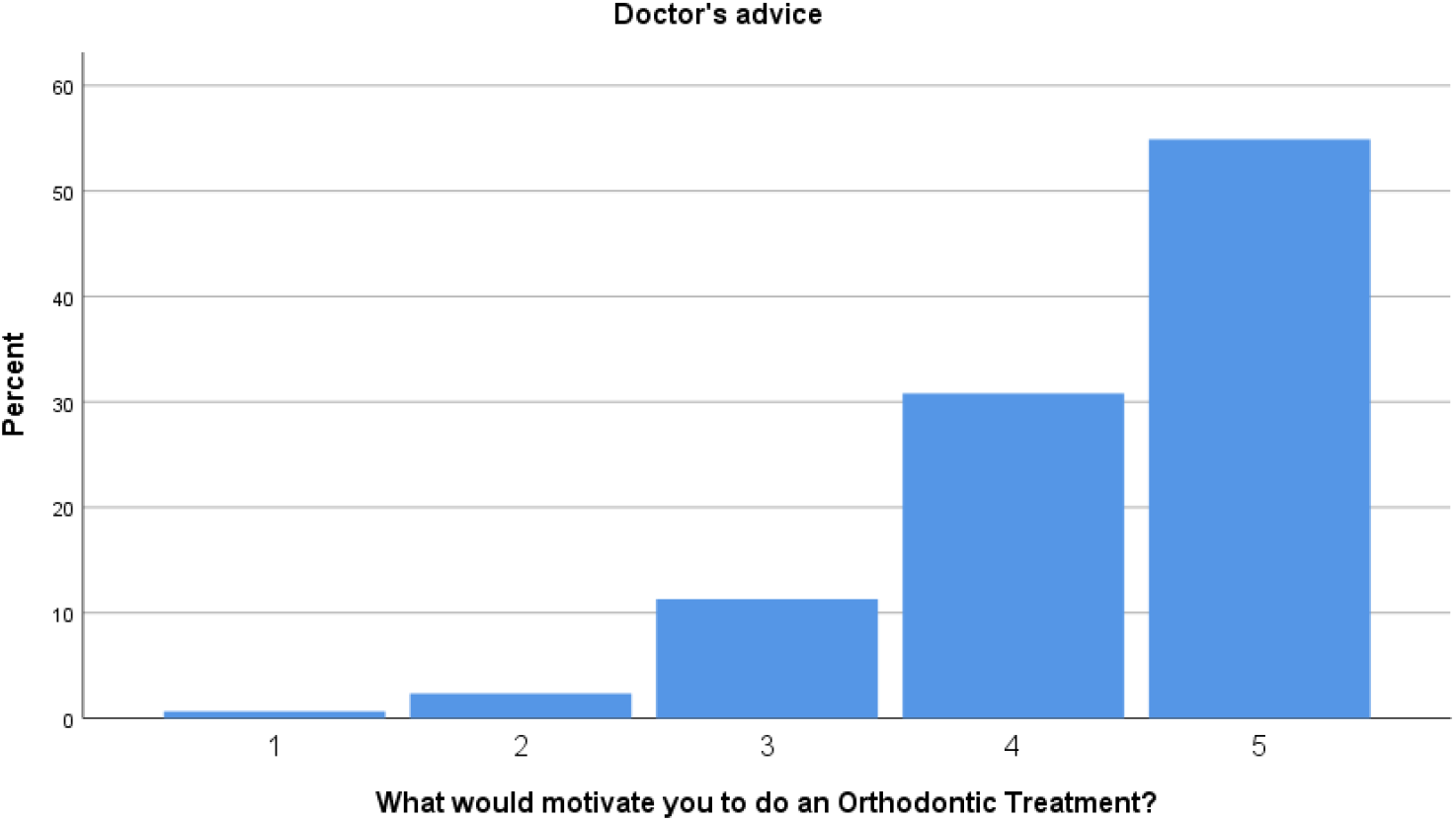
What would motivate you to do an Orthodontic Treatment – Doctor’s Advice

**Image 4.**
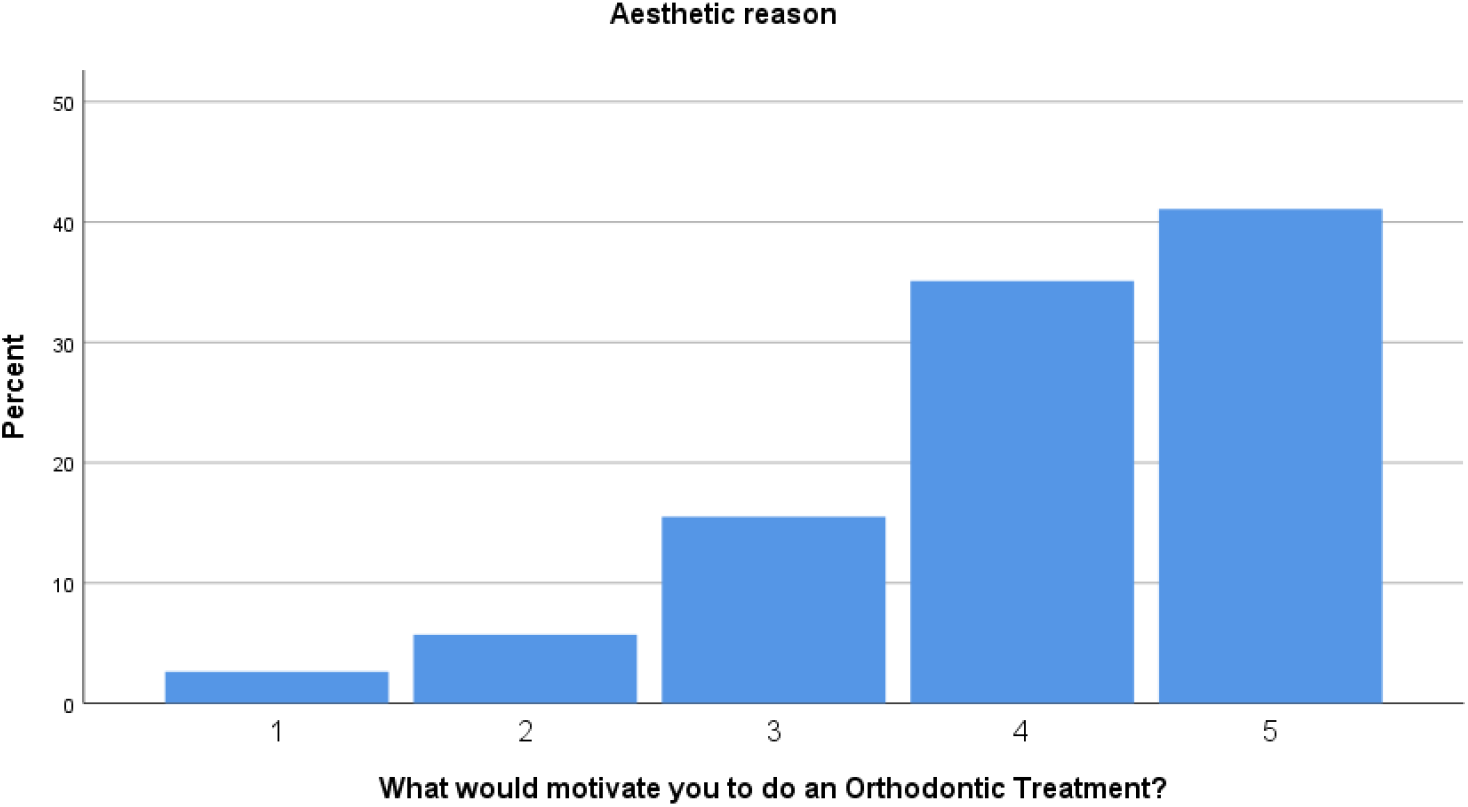
What would motivate you to do an Orthodontic Treatment – Aesthetic Reason

**Image 5.**
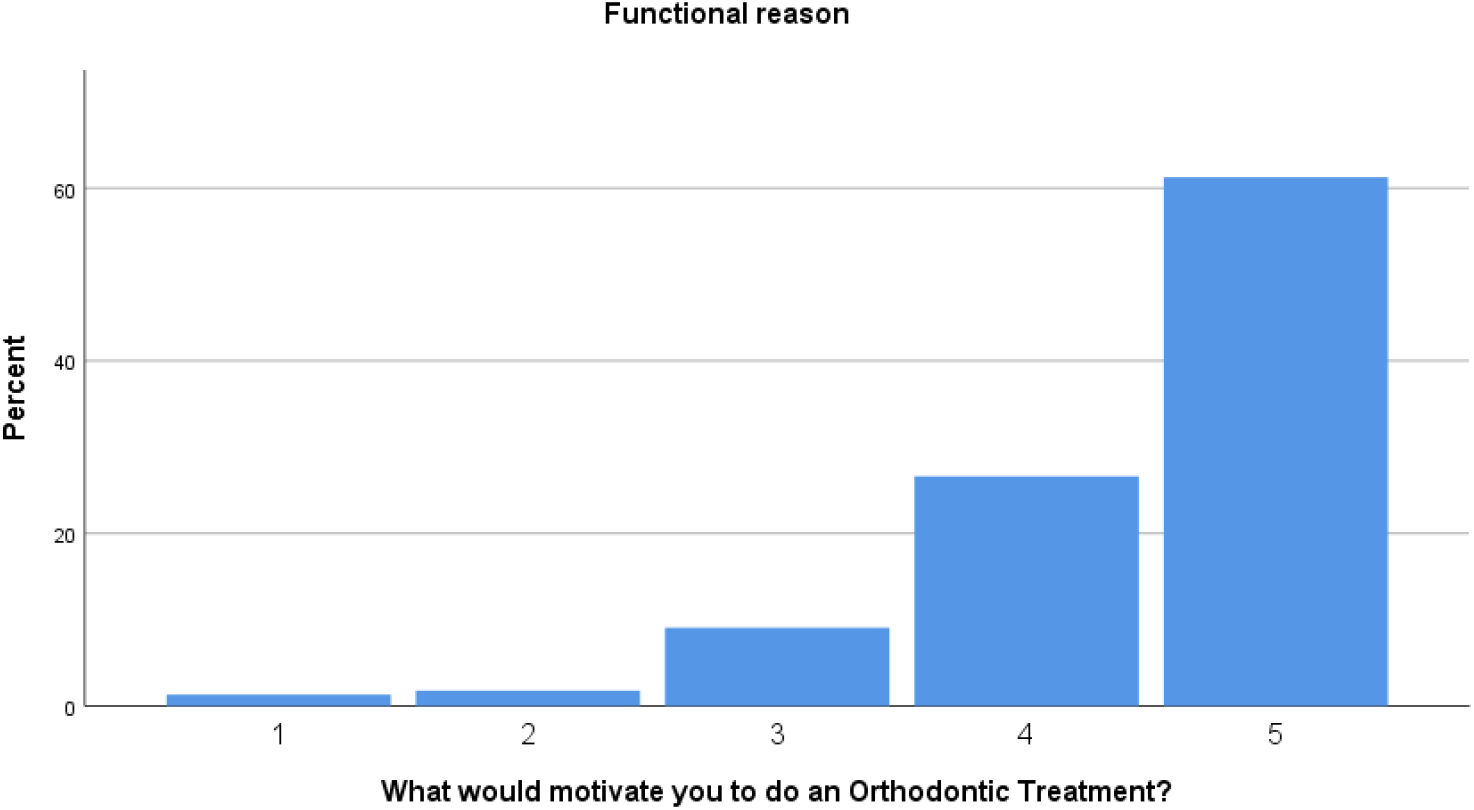
What would motivate you to do an Orthodontic Treatment – Functional Reason

**Image 6.**
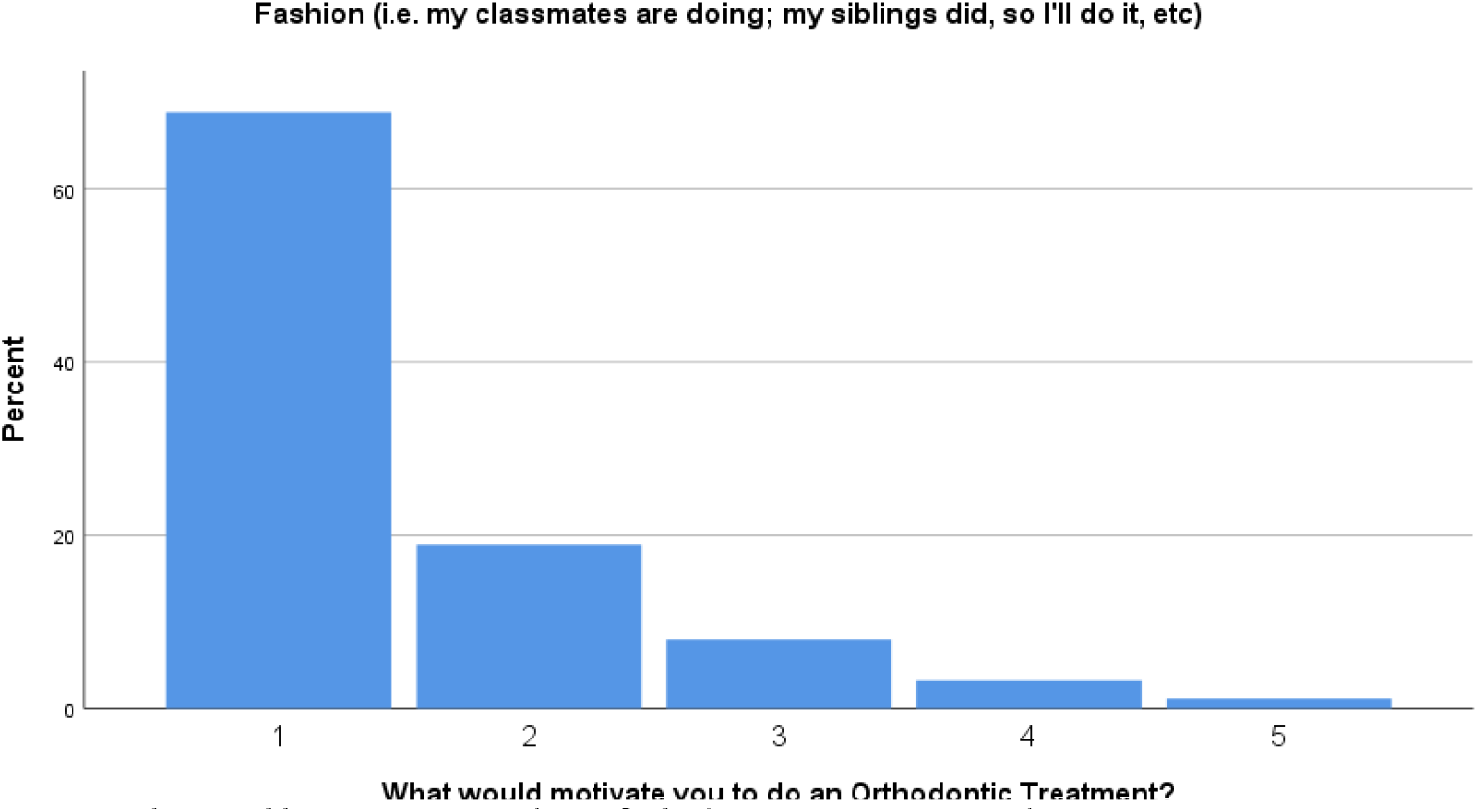
What would motivate you to do an Orthodontic Treatment – Fashion

### Comparing Nationalities and Universities

#### Students – Portuguese *versus* Romanian

In order to compare, it was used the T-Test for Independent Samples with a margin of error alpha of 5%. The following results were found:

**Table I.**
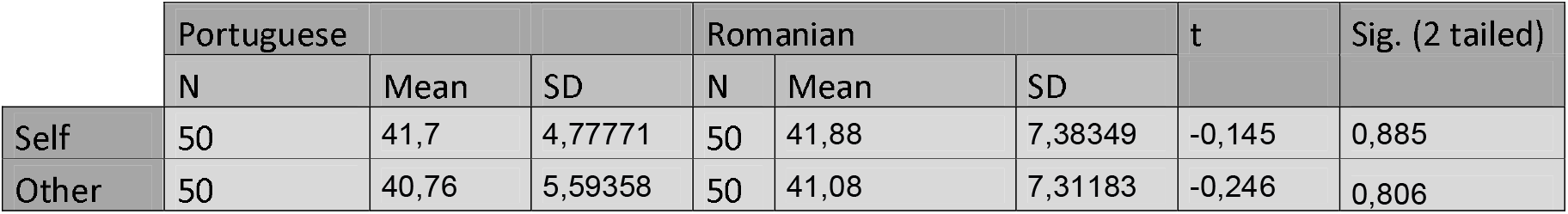
Results of comparing the means of Portuguese and Romanian Students

|  | Portuguese |  |  | Romanian |  |  | t | Sig. (2 tailed) |
| --- | --- | --- | --- | --- | --- | --- | --- | --- |
|  | N | Mean | SD | N | Mean | SD |  |  |
| Self | 50 | 41,7 | 4,77771 | 50 | 41,88 | 7,38349 | -0,145 | 0,885 |
| Other | 50 | 40,76 | 5,59358 | 50 | 41,08 | 7,31183 | -0,246 | 0,806 |

**Image 7.**
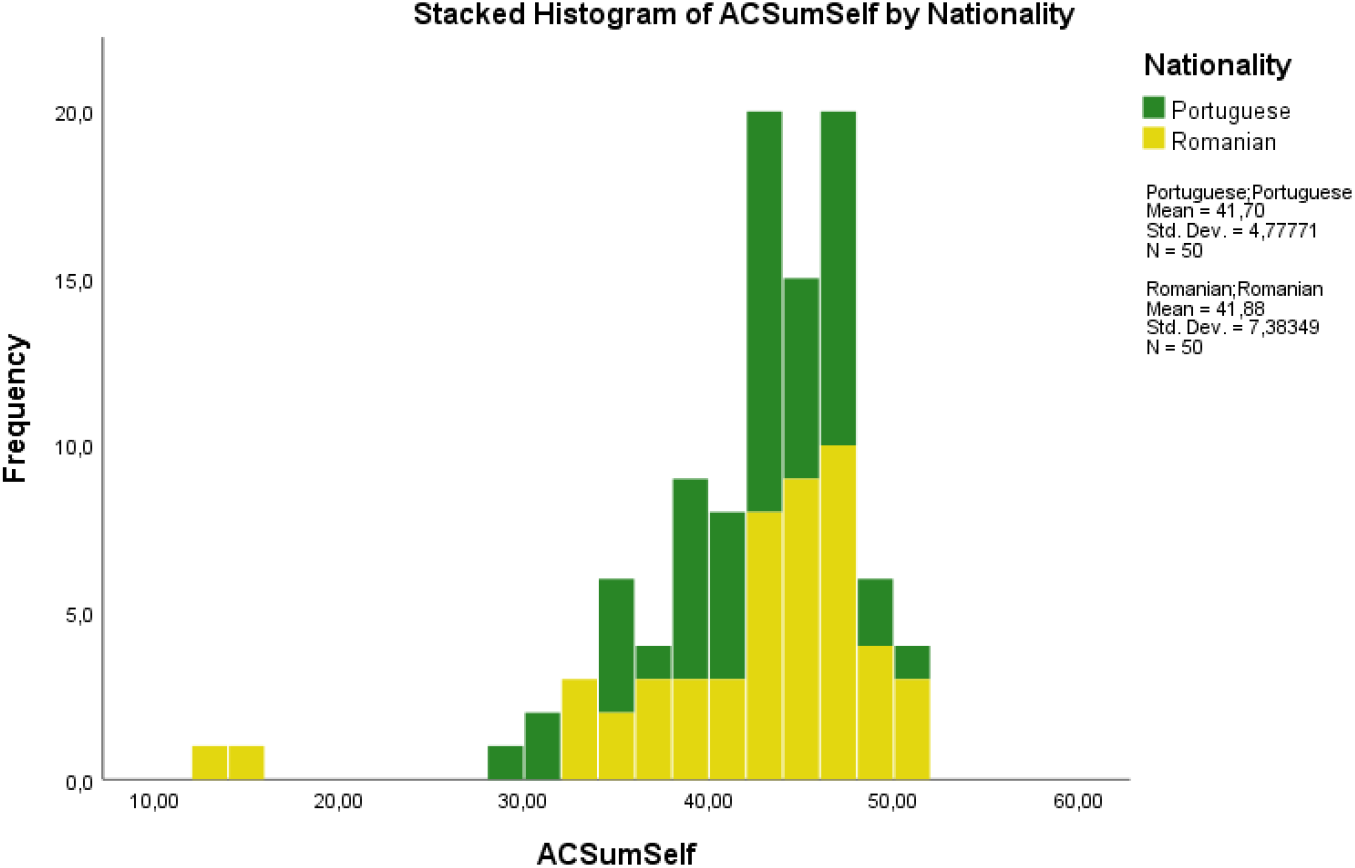
Histogram of the comparison of Portuguese and Romanian Students – Self-Perception

**Image 8.**
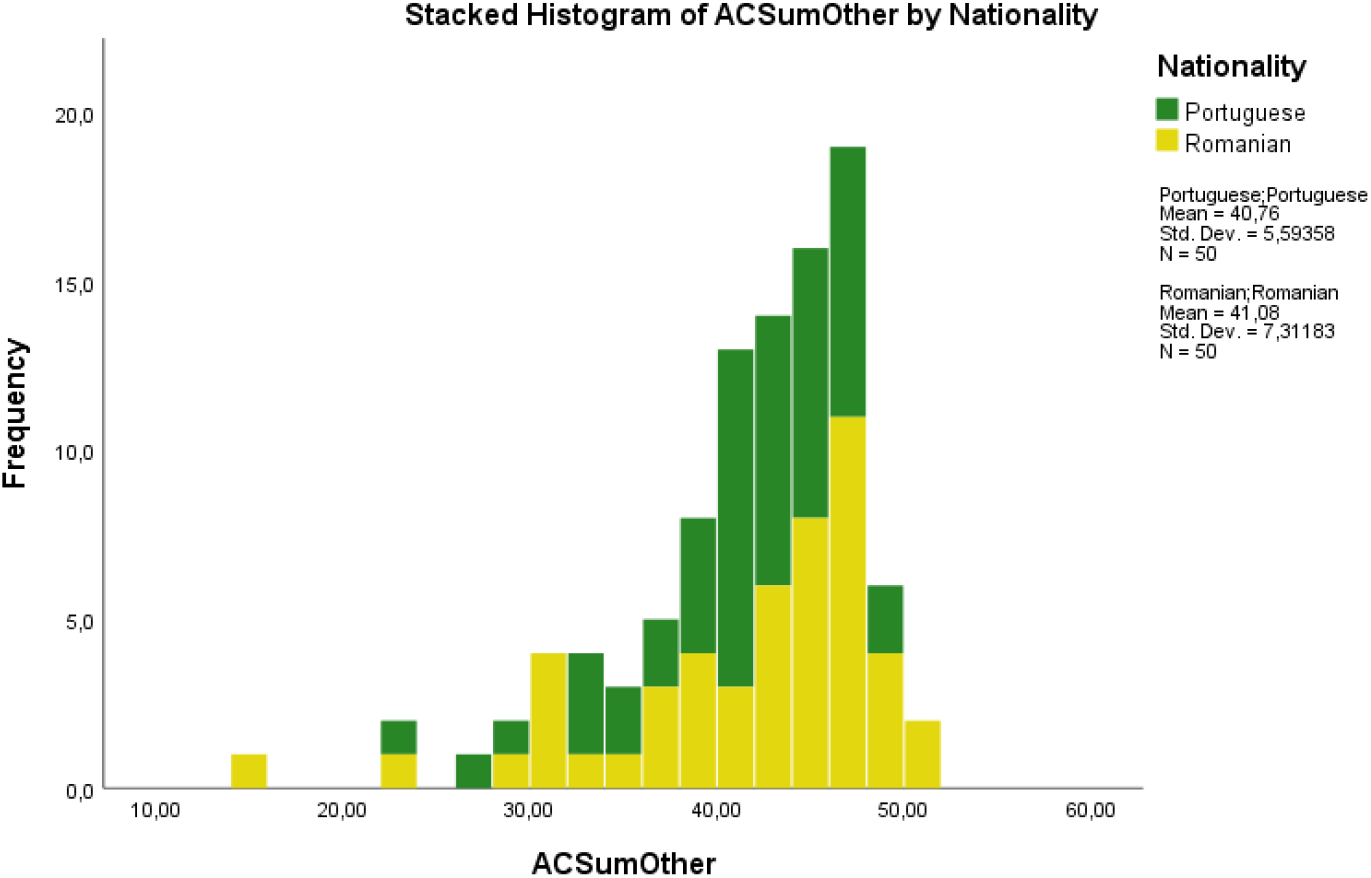
Histogram of the comparison of Portuguese and Romanian Students – Others Perception

The results show no statistical differences between Portuguese and Romanian Students’ OES comparison for Self-Perception nor for Others.

Dentistry Students – Portuguese *versus* Romanian

The following results were obtained:

**Table II.** Results of comparing the means of Portuguese and Romanian Dentistry Students

|  | Dentistry Students |  |  |  |  |  |  |  |
| --- | --- | --- | --- | --- | --- | --- | --- | --- |
|  | Portuguese |  |  | Romanian |  |  | t | Sig. (2 tailed) |
|  | N | Mean | SD | N | Mean | SD |  |  |
| Self | 35 | 42,3714 | 4,06626 | 35 | 42,5714 | 6,33053 | 0,058 | 0,954 |
| Other | 35 | 41,8571 | 5,16289 | 35 | 41,7714 | 7,09634 | -0,157 | 0,876 |

**Image 9.**
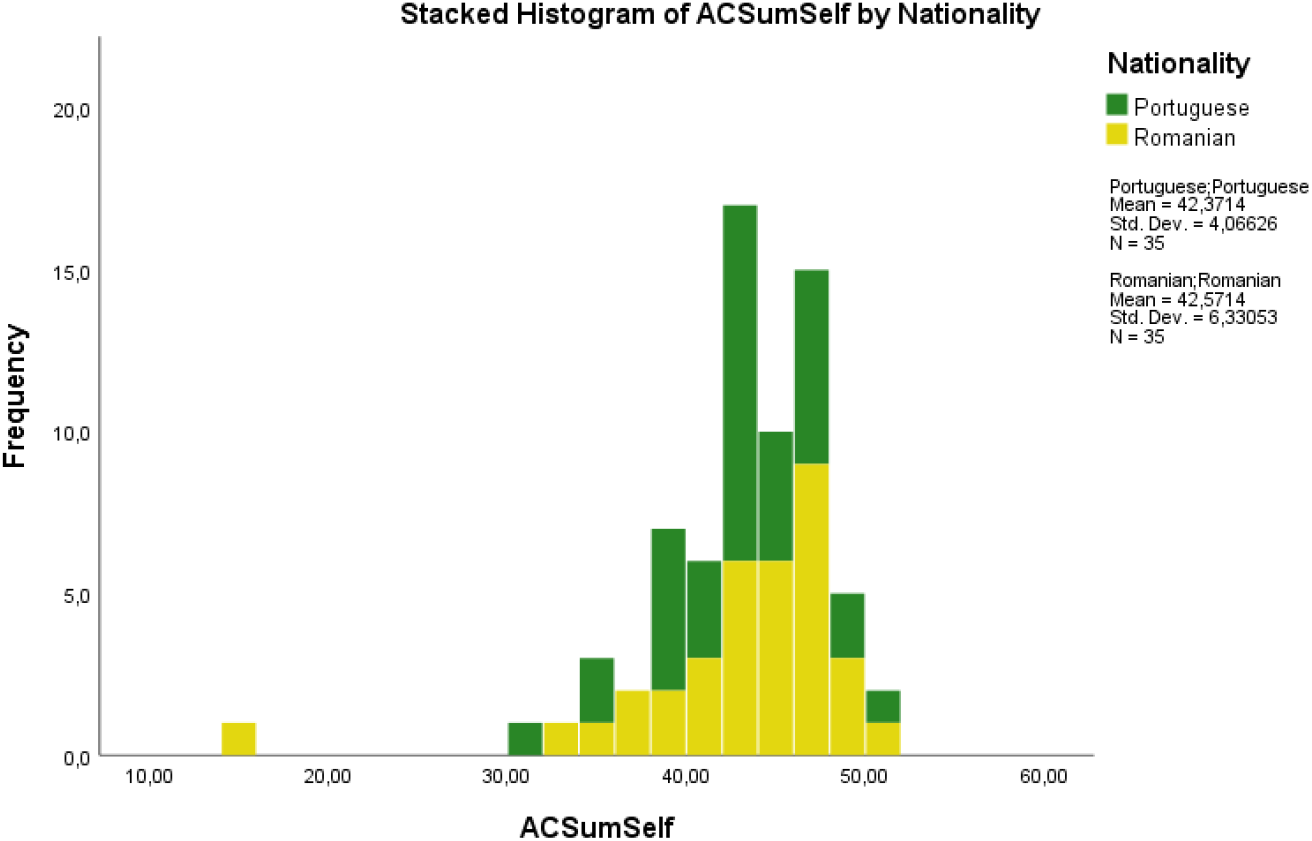
Histogram of the comparison of Portuguese and Romanian Dentistry Students – Self-Perception

**Image 10.**
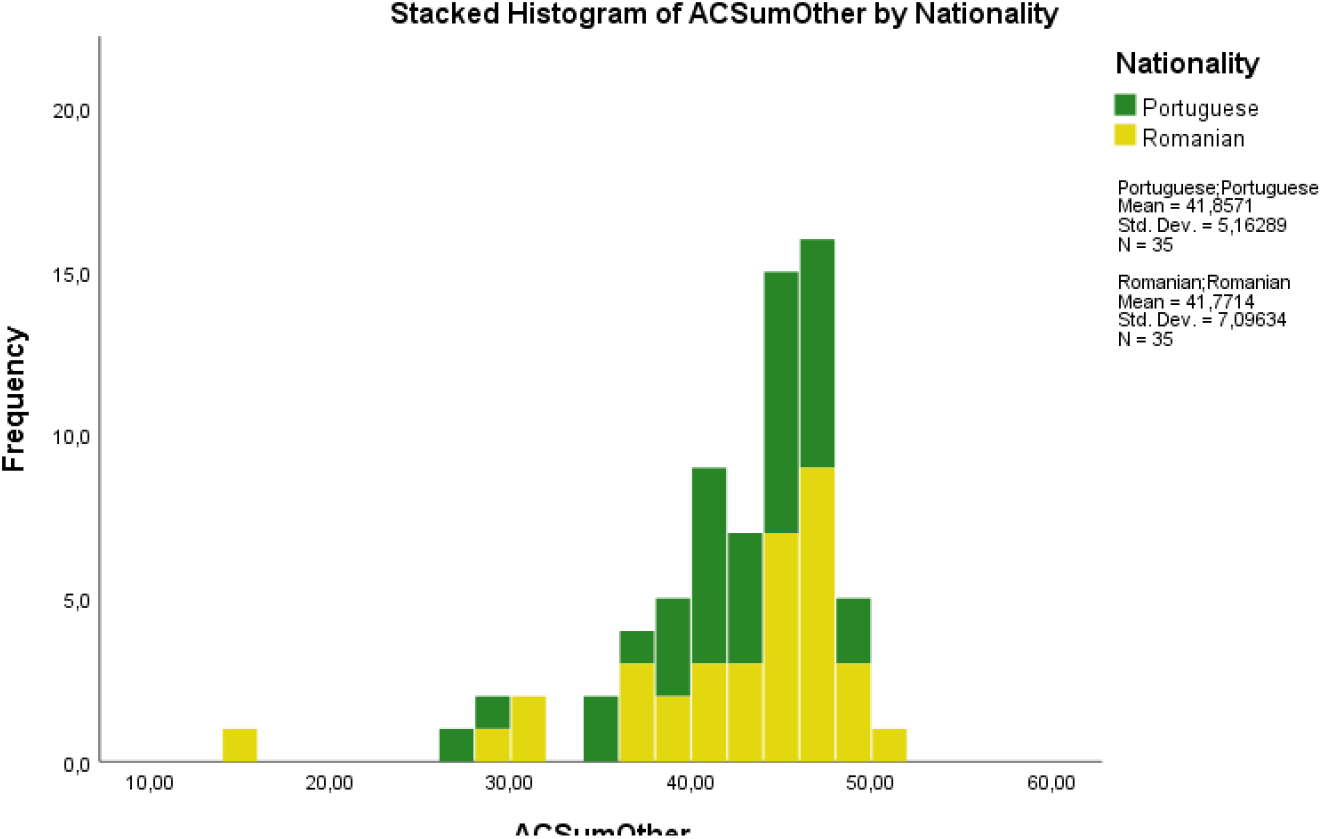
Histogram of the comparison of Portuguese and Romanian Dentistry Students – Others Perception

Dentistry students show no statistical differences regardless of their nationality.

UMF Dentistry Students – Romanian Students *versus* International

**Table III.** Results of comparing the means of Romanian and International Dentistry Students of UMF

|  | UMF Dentistry Students |  |  |  |  |  |  |  |
| --- | --- | --- | --- | --- | --- | --- | --- | --- |
|  | Romanian |  |  | International |  |  | t | Sig. (2 tailed) |
|  | N | Mean | SD | N | Mean | SD |  |  |
| Self | 35 | 42,5714 | 6,33053 | 58 | 41,4483 | 6,79828 | 0,792 | 0,431 |
| Other | 35 | 41,7714 | 7,09634 | 58 | 40,9828 | 5,99851 | 0,573 | 0,568 |

**Image 11.**
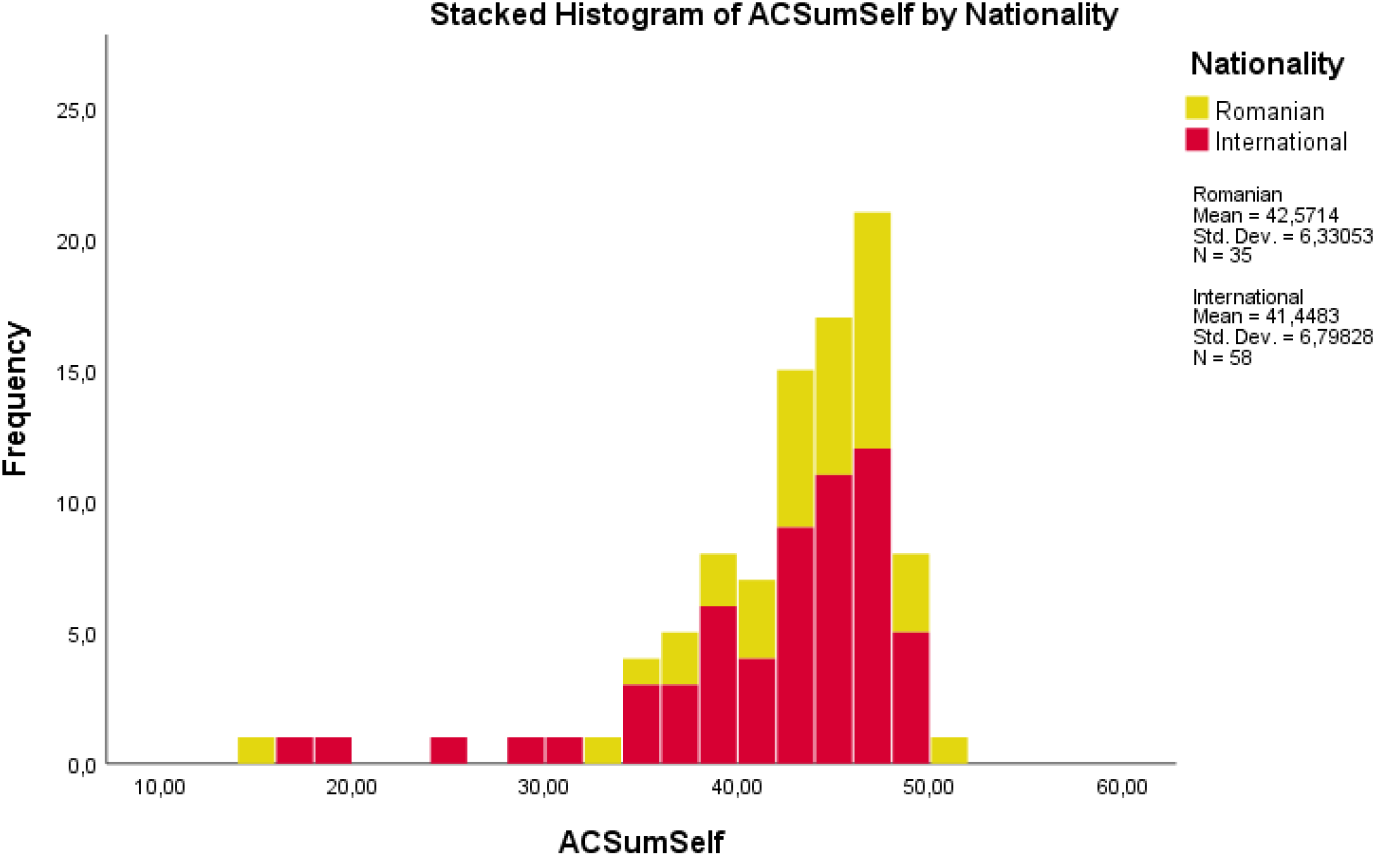
Histogram of the comparison of Romanian and International Dentistry Students of UMF – Self-Perception

**Image 12.**
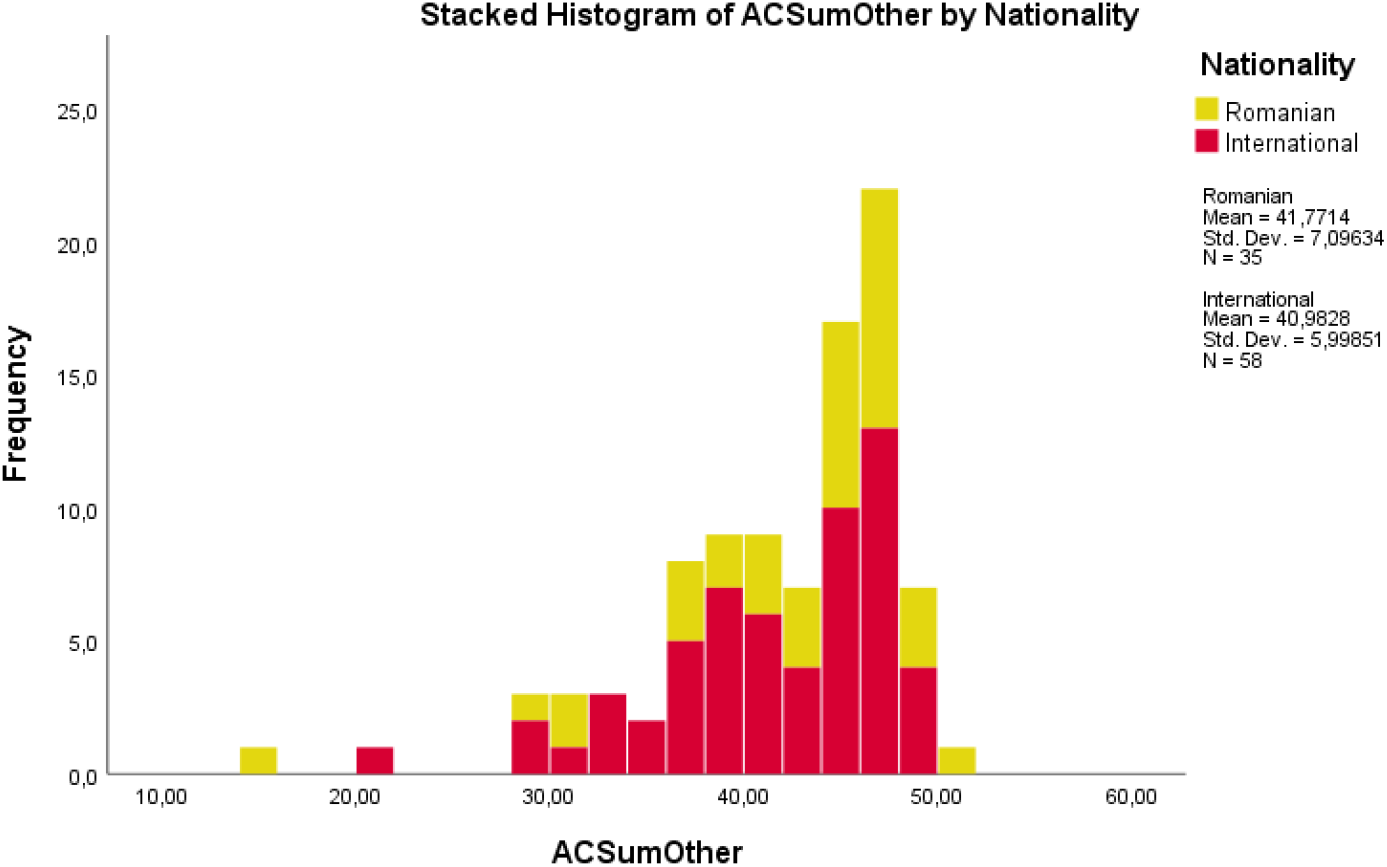
Histogram of the comparison of Romanian and International Dentistry Students of UMF – Others Perception

There are no statistical differences between International Students in Romania and Romanian Students.

### Comparing the generality of University Students

#### Academic Degree

The results obtained were:

**Table IV.**
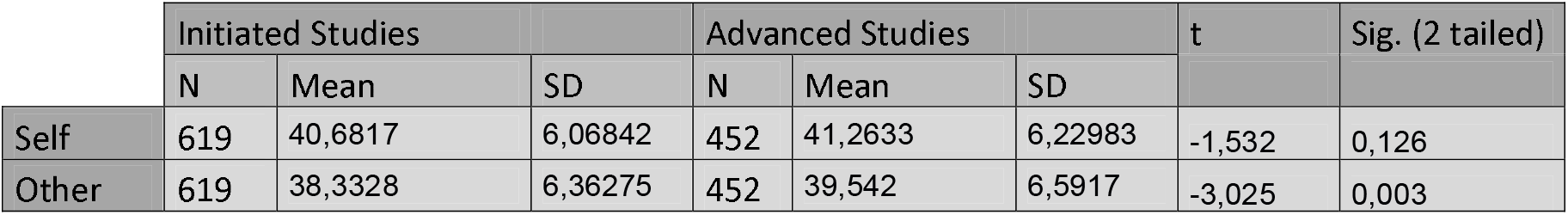
Results of comparing the means of Initiated and Advanced Studies in generality

|  | Initiated Studies |  |  | Advanced Studies |  |  | t | Sig. (2 tailed) |
| --- | --- | --- | --- | --- | --- | --- | --- | --- |
|  | N | Mean | SD | N | Mean | SD |  |  |
| Self | 619 | 40,6817 | 6,06842 | 452 | 41,2633 | 6,22983 | -1,532 | 0,126 |
| Other | 619 | 38,3328 | 6,36275 | 452 | 39,542 | 6,5917 | -3,025 | 0,003 |

**Image 13.**
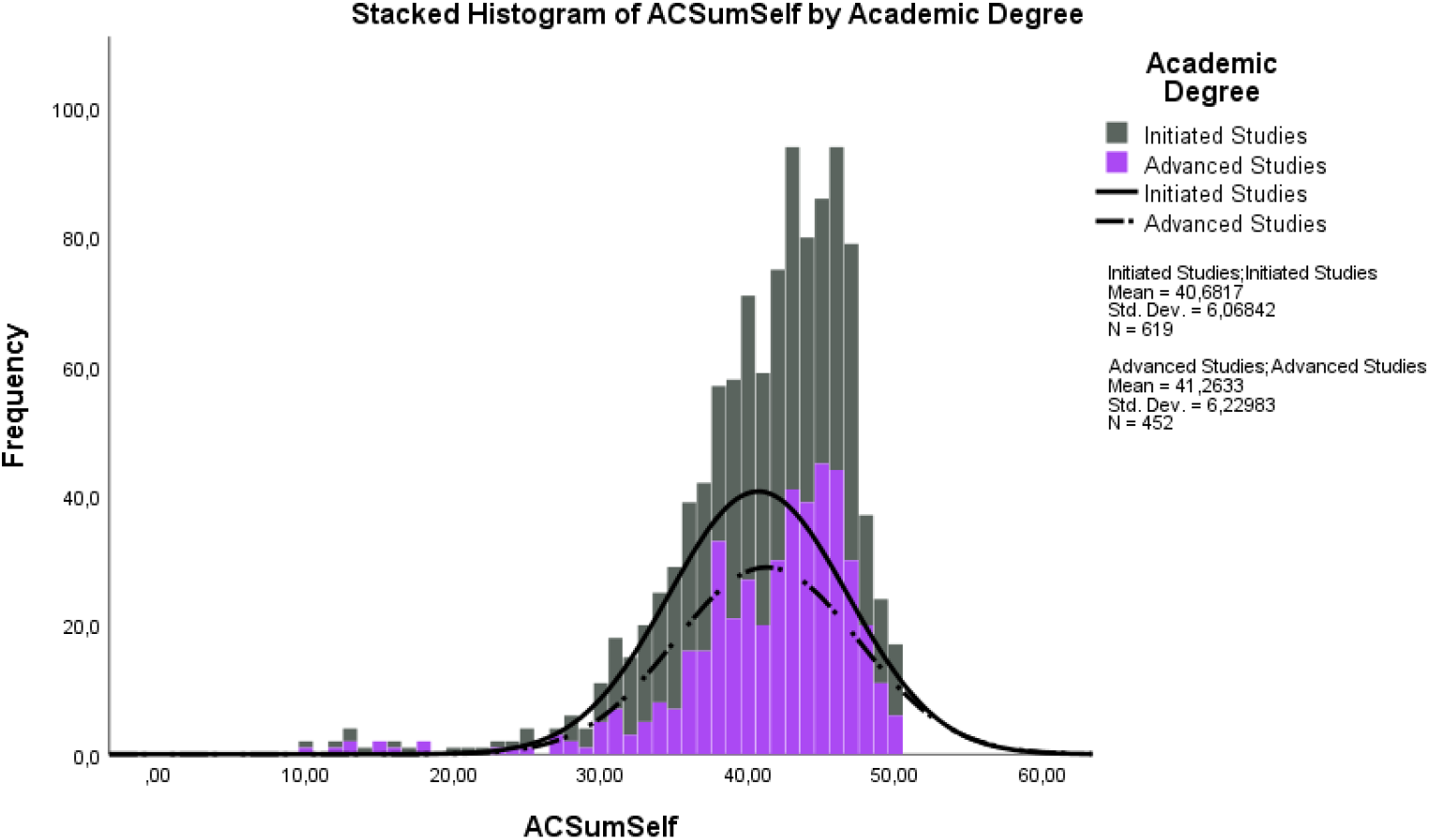
Histogram of the comparison of Initiated and Advanced Students – Self-Perception

**Image 14.**
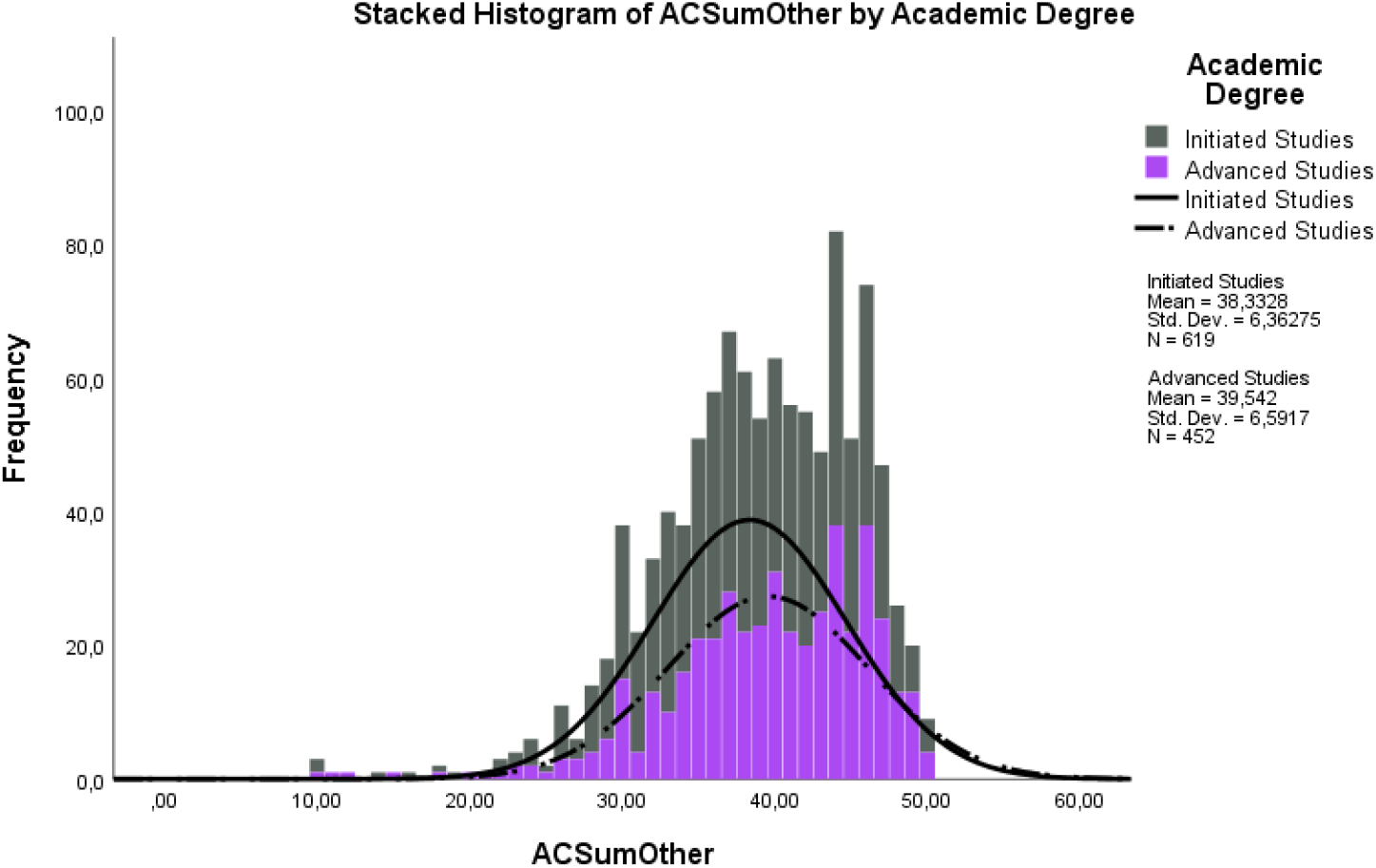
Histogram of the comparison of Initiated and Advanced Students – Others Perception

There are statistically significant differences between OES means of Initiated Students (M=38,3328; SD=6,36275) and Advanced Students (M=39,542; SD=6,5917) for Others Perception. (t=-3,025; p=0,05). No differences for the Self.

Initiated Students vs. Advanced Students among different Courses of Studies

**Table V.**
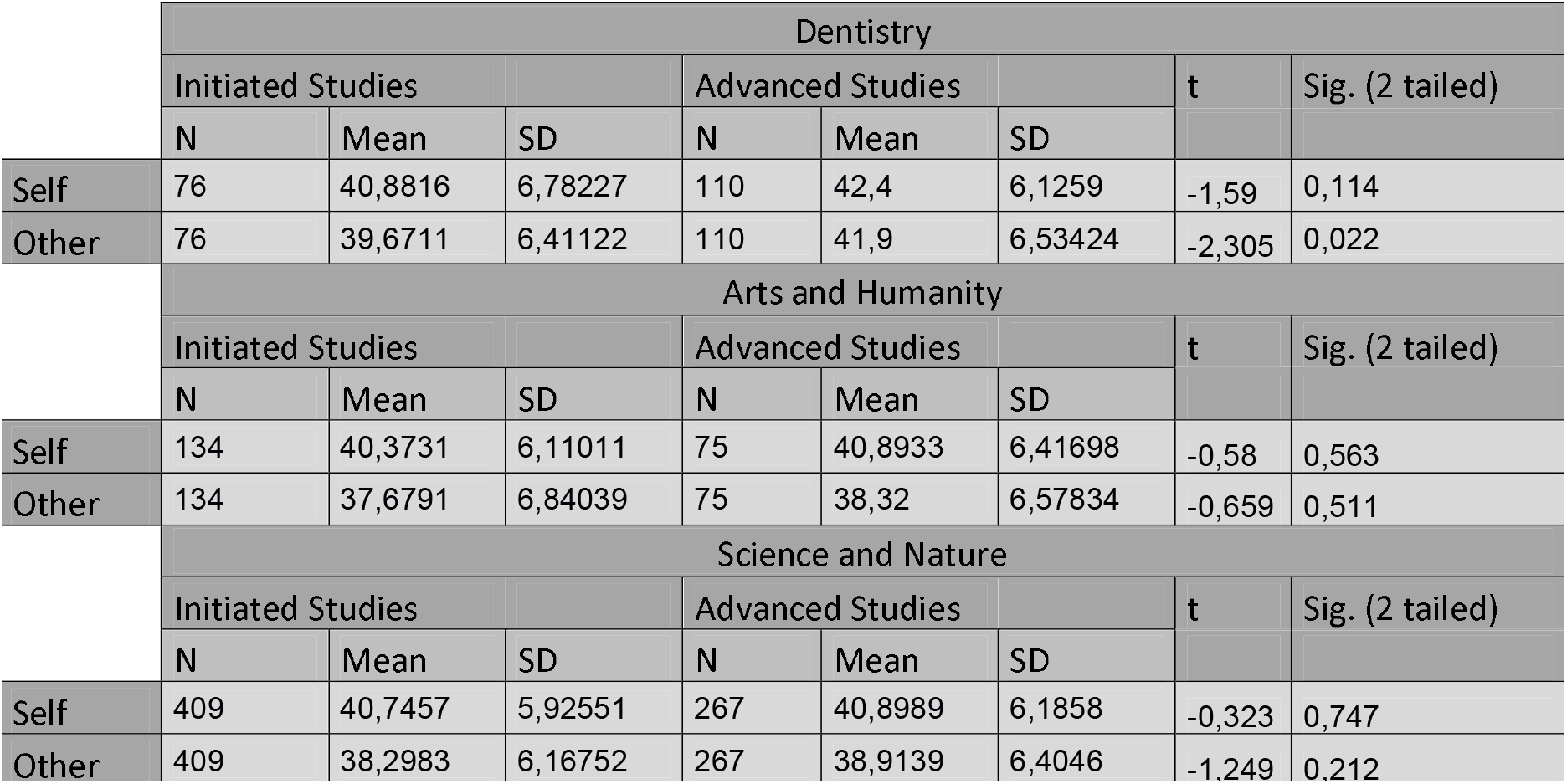
Results of comparing the means of Initiated and Advanced Studies among Courses of Studies

There are statistically significant differences in OES means:

- Other:
  1. Dentistry:
    - Initiated Students – M=39,6711; SD=6,4122
    - Advanced Students – M=41,9; SD=6,53424
      - t=-2,305; p=0,05

No other comparison between OES means showed statistically significant differences.

Arts and Humanity vs. Science and Nature

**Table VI.**
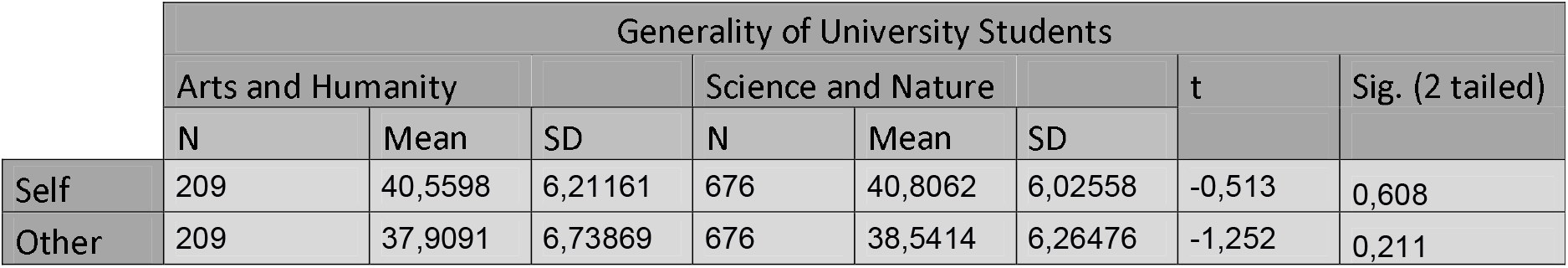
Results of comparing the means of Arts and Humanity and Science and Nature

Dentistry vs. Non-Dentistry Studies

**Table VII.**
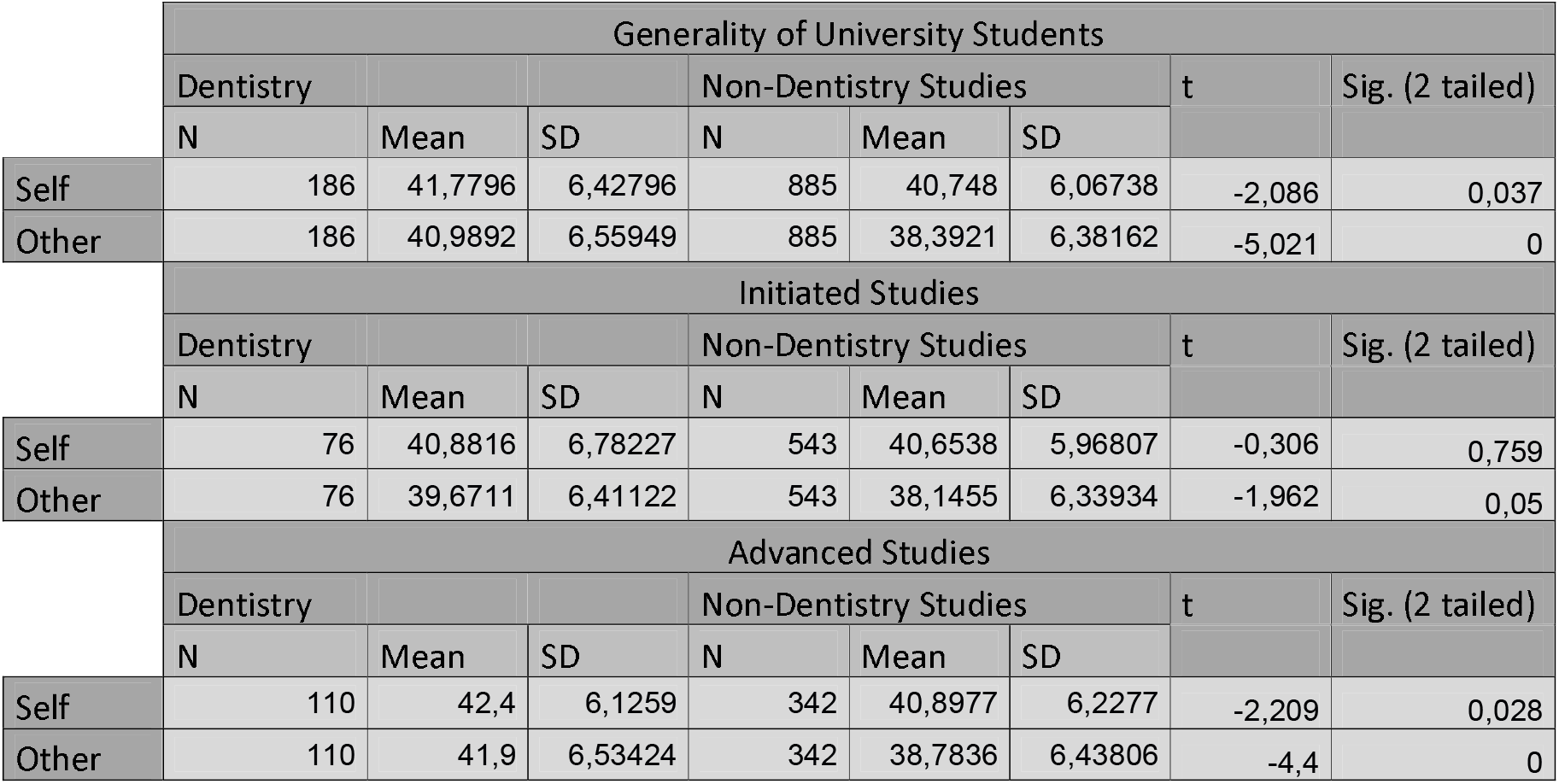
Results of comparing the means of Courses of Studies in generality and among Academic Degree

There are statistically significant differences of OES means:

- Self:
  1. Generality:
    - Dentistry Students – M=41,7796; SD= 6,42796
    - Non-Dentistry Students – M=40,748; SD= 6,06738
      - t=-2,086; p=0,05
  2. Advanced Students:
    - AS Dentistry – M=42,4; SD=6,1259
    - AS Non-Dentistry Students – M=40,8977; SD=6,2277
      - t=-2,209; p=0,05
- Other:
  1. Generality:
    - Dentistry Students – M=40,9892; SD=6,55949
    - Non-Dentistry Students – M=38,3921; SD=6,38162
      - t=-5,021; p=0,05
  2. Advanced Students:
    - AS Dentistry – M=41,9; SD=6,53424
    - AS Non-Dentistry Students – M=38,7836; SD=6,43806
      - t=-4,4; p=0,05

OES Self-Perception vs. Others Perceptions

**Table VIII.**
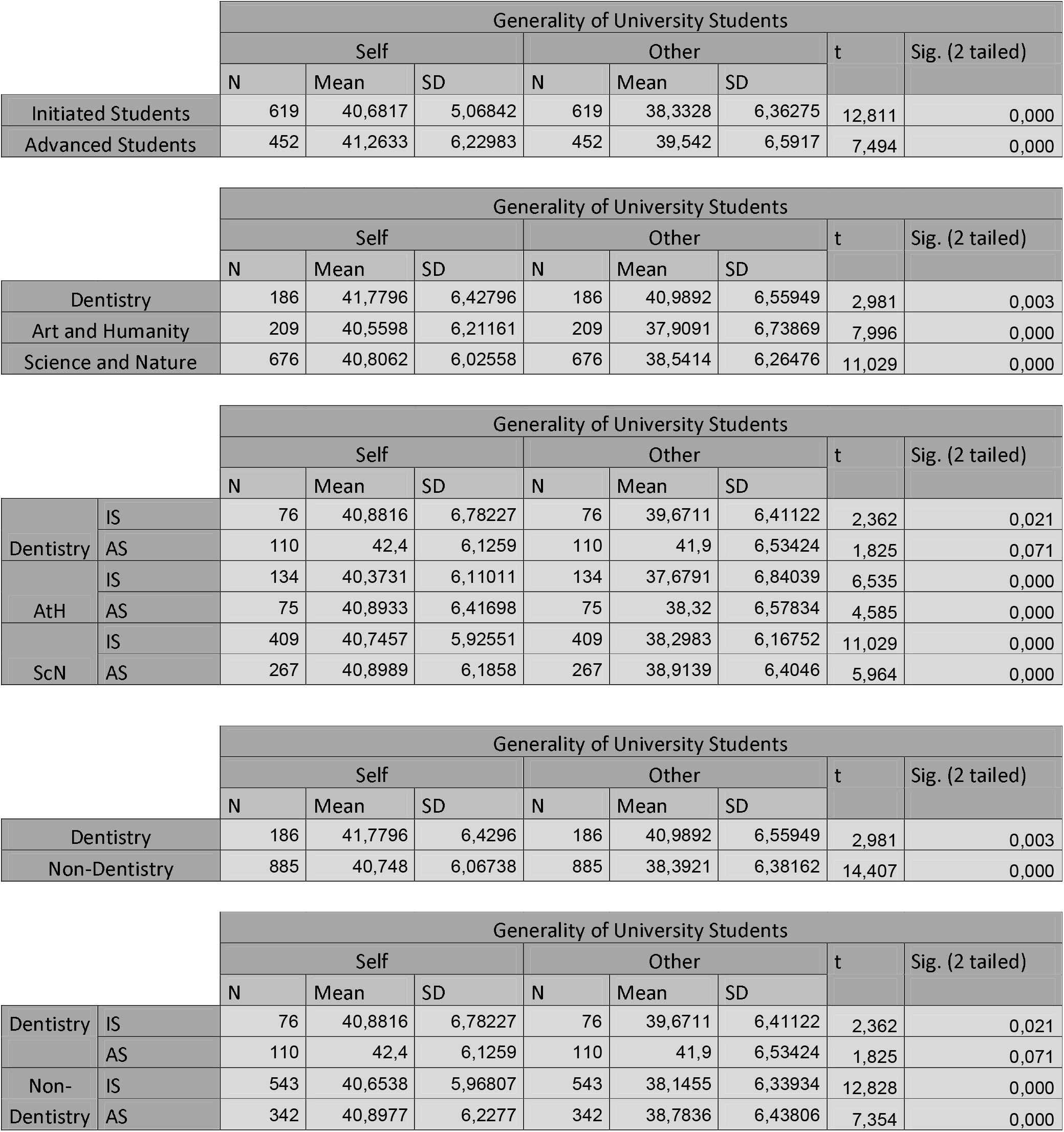
Results of comparing the means of Self-Perception and Perception of Others among all Groups

There are no statistically significant differences between the means of Self OES and Other OES in Advanced Students of Dentistry.

All other differences are statistically significant.

## Discussion

### Sample

Older participants, born before 1980, were a few outliers, not considered for this study in order to keep the cohesion of the age groups among the students.

The decision of grouping the participants with the described criteria of Initiated and Advanced Students was made due to the fact that a 4^th^ year Dentistry student in UP or UMF does not have the clinical experience that a student of the last or 5^th^ years has. The same is applied to a student of the 3^rd^ year in Humanistic Studies or 4^th^ year in Natural Studies, being, their penultimate year. That is, their thinking maturity isn’t always as focused as a finalist, inevitably on the verge of facing the employment world and, sometimes, keeping the development of their studies in the process.

The group Dentistry was composed of students from FMDUP and UMF.

The participants from courses of studies such as Engineering, Natural Sciences, Economics, Health, Sports, or Pharmaceutical Studies were considered as ScN, whilst Arts, Architecture, Literature, Law, History, or Humanistic Studies were considered as AtH. This clustering was based on the backbone of the respective studies. Science and Nature students are instigated to think or to perform at a more objective standard, following protocols to achieve an expected outcome. They are often dwelling on matters that have already been studied. On the other hand, Arts and Humanity Studies have their baseline of fundamental rules or principles and with different arguments, points of view, interpretations, or expressions one can achieve a great variety of subjective results for the same challenging situation.

Dentistry is an intriguing mixture of Art and Science, both lines of thinking. It is mostly objective and standardized in the matters of oral health, however, restoration, rehabilitating, or improving the aesthetical appearance of a patient is a very subjective field. There are plenty of approaches that may focus more on surgery, prosthetics, or orthodontics, depending on the patient’s health and wishes. More specifically, orthodontists are a combination of the engineer and architect of the building in construction, which is the oral cavity of a growing child, for example. They act on an attractive appearance and competent function. ^(13, 19, 20)^

### Statistical Analysis of OES

The OES was created to characterize the respondent, without segregation or a competence label.

The ratings, from 1 to 5, for each of the 10 pictures of the AC were summed resulting in a single rating (per observer) with a score ranging from 10 to 50. Used for the Self-Perception and another for the Perception of Others, to find out if there are significant differences between the participants’ mean OES, being judged by their subjective opinion, and not by their competence.

### The Index of Orthodontic Treatment Need

It is important to note that many participants considered the Aesthetic Component of the Index of Orthodontic Treatment Need as poor in image quality. The pictures are as recent as 1989, at the most, which builds a gap of 31 years to the present day, meaning that the quality is not as good as what we can now achieve. The main problem is a big discrepancy between the first Clinical Cases presented and the last, that is, some participants stated that they were rating a low score and then all of a sudden were rating the maximum necessity level, lacking a harmonic decrease in the cases appearance and health. Many photographs are borderline unanimous, leaving little room for subjective thinking and divergence of perceptions.

Just as Borges (2015) ^(17)^ stated, it’s true that dental aesthetics differ from person to person and possibly from nation to nation, although it’s verified that it is not expressive in the majority of cases. It is added that one disadvantage is the inclusion of frontal intraoral pictures only, disregarding anteroposterior planes, malocclusions of overbite, open bite, agenesis, supernumerary teeth, and other kinds, implying that the AC may not reflect the true variety of opinions.

Brook et al. (1989) ^(15)^ referred, that the absence of side views may have biased some opinions on the cases that for a layperson may not be too displeasing. On the other hand, the original inclusion of both front and side views led to many inconsistencies.

While these perspectives are extremely pertinent for Clinicians and Advanced Students of Dentistry, they can be overwhelming for the Initiated Students and the groups of Non-Dentistry Studies, who would play judge on arranging a new set of pictures, if it were to update the IOTN.

An index is desired to be as precise as possible, however, every patient is different. When left with only a few options for discussion, opinions will be biased. One can never discard the little nuances that are not so determinant to the diagnostic but undeniably exist. However, the IOTN is the most used index and it is validated, therefore, in the logistic impossibility to create and validate an index of this nature in a timely manner, it is preferable to use a validated index than to base the study on a non-validated tool.

### University Students and Orthodontic Treatment outcome

There are many studies on comparisons between the perception of many social groups, including Dentistry Students, to assess if the population perceives a bigger or smaller need for treatment or if it has any psychosocial impact. Nonetheless, this research seems to be pioneer in assessing the way of thinking of Students of different Academical areas. Every group has its own trend of studying and thinking, but more similar rather than different, regarding the aesthetics of the oral cavity.

The statistical differences between the means of all AS and IS could be a small sign of the unassertiveness of the latter. There is a common occurrence in the early years of University, with a tendency to change when growing up.

The lack of statistical differences between IS in Dentistry and IS in Non-Dentistry Studies and between the areas of AtH and ScN is not surprising. It may be because the respondents are not so learned in oral matters and, therefore, do not feel ready to be critical enough to judge its own oral cavity, much less other person’s. This considering the fact that throughout the many comparisons, the mean Perception of the Other is always rated with a lower mean OES than the Self.

However, when it comes to the decision of the AS in Dentistry, even though the difference of Self OES mean is not statistically significant to an IS in Dentistry, it is still considerable. Unsurprisingly, this group registers the biggest mean measure of OES for the Perception of Others and of the Self and is, most certainly, because of the confidence felt on the knowledge acquired in this stage of the graduation, closer to the Employment.

It’s interesting to notice that these are contrary to the results found by Borges (2015) ^(17)^. She verified that first year Students in Dentistry would give more importance to the OT. The participants from the Polish University she studied rated higher necessity for OT than students from FMDUP and she did not find statistical differences between the perception of the Self and Others. It is important to add, that the methodology used is not the same used in the present study.

As suspected at the beginning of the initial idea of the study, when comparing the respective OES, the participants of the Arts and Humanity area have a significance of 56,3% and 51,1% between Advanced and Initiated Studies, that is, their range of means is not statistically meaningful. This shows precisely that the logical process of decision is very little consistent and more a matter of sensitivity and interpretation.

The Science and Nature area participants show a significance of 74,7% for the Perception of the Self, between Advanced and Initiated Students and 21,2% when regarding Other people, which means a statistical “disagreement” of 78,8%, signs of a more fixed position. A less subjective opinion on what is beautiful and what is not. The same way of thinking as the likes of Dentistry Students, which is obviously part of the Science and Nature area and play the utmost important role of the Professional opinion in the present study.

These assumptions are statistically supported and are verified by the rule of thumb nonetheless, they should be taken as a support, as the results of the method used find differences, not extrapolations and it would be very interesting to test again with a more incisive method.

The challenge and a possible ongoing project for this study could be to create a new version of the IOTN addressing all the suggestions that have been made and with new and better-quality pictures. Could also be included a Psychological Component along with the existing ones, with interest to psychosocial effects on NHS patients who went through OT. It should be studied in two different times – Before OT and After OT, ideally, including more stages – During OT and up to 10 years after OT. ^(6-^8, 14, 21^)^

## Conclusion

For the studied population of students, the following is concluded:

There is an agreement in Doctor’s Advice, Aesthetic and Functional Reasons as the most prominent motivations to look for Orthodontic Treatment, whereas Fashion is the least weighing factor.

OES for Perception of Others is influenced by academic degree, specifically in Dentistry students

OES for Self-Perception and for Perception of Others is similar only for Advanced Dentistry students

OES is influenced by Dentistry studies, specifically in advanced students

OES is not influenced by the country students grew up nor the country they are graduating

OES seems not to be influenced by the so-called subjectivity or objectivity of undergraduate studies, in the meaning of comparing education in Arts and Humanity with Science and Nature.

## Supporting information

Annex Section

Transparency Declaration

Data Sharing Statement

CONSORT guidelines

Author Disclosure Alves

Author Disclosure Mesaros

Author Disclosure Ponces

Author Disclosure Pollmann

## Data Availability

All data is available when requested via email to

## Notes

Funding Source: None

Conflict of Interest: None

### Competing Interest Statement

The authors have declared no competing interest.

### Funding Statement

No funding was supported

### Author Declarations

The inquiry was approved by the Ethics Commission of FMDUP and the UP's Data Protection Office, being the verdict valid to the Dean's office of UMF (annexes section - The original Portuguese document was issued on 03/01/2020 and is available upon request. The English translation was issued on 17/12/2020). The study was part of a Master's Degree Research Thesis for the University of Porto in collaboration with the University of Medicine and Pharmacy of Cluj-Napoca, therefore before any chance of being published by MedRXiv, it was already validated and authorized by both institutions and their respective Ethics Commission/Dean's Office. All participants collaborated voluntarily and anonymously.

### Summary of Updates

Sub Title topics were added to the Abstract. The Distribution/Reuse Options have been revised. Anyone can share this material, provided it remains unaltered in any way, this is not done for commercial purposes, and the original authors are credited and cited.

