## Supplementary material for "Aesthetic evaluation of the need for orthodontic treatment – Perception among university students": Annex Section

### Annexes Section

Annex 1

Inquiry used for the study

### Inquiry for orthodontic treatment need

The following inquiry is part of a study in Dentistry for Luís Alves's Master's Degree Final Report "Aesthetic evaluation of the need for orthodontic treatment - Perception among University students/Necessidade estética de tratamento ortodôntico - Perceção de estudantes universitários", as a student from University of Porto Faculty of Dentistry (FMDUP).
The images used were taken from the “Aesthetics Component” from the Index of Orthdontic Treatment Need (IOTN).
The research will strictly be used for academic and scientific purposes and aims to compare different opinions regarding malocclusions (imperfect positioning of the teeth when the jaws are closed). Your personal and physical integrity will not be exposed to any risk. You will not receive any kind of payment or monetary gratification for participating.
It's anonymous. No identification shall be left on any page, in order to guarantee confidentiality in every response. The data will never be published as a way to identify you.
By answering the following questions and proceed to fulfill this inquiry, you agree to be part of this study and consent to be included in the database, on which the results, and following conclusion, will be based upon. You are free to choose not to answer, or give up at any moment under no penalty or cost.
This inquiry was approved by the University of Porto's Data Protection Office and the Faculty of Dentistry's Ethics Comission.
Please read carefully every page and the clinical situation presented.
Fill in the blanks according to your opinion and answer every question with all your honesty.
Thank you for your collaboration and remember that there are no correct/incorrect answers, only opinions that will be collected as information to be treated anonymously respecting all confidentiality guidelines.
* Required

1. Year of Birth *

*Mark only one oval.*

- ( ) 1990
- ( ) 1991
- ( ) 1992
- ( ) 1993
- ( ) 1994
- ( ) 1995
- ( ) 1996
- ( ) 1997
- ( ) 1998
- ( ) 1999
- ( ) 2000
- ( ) 2001
- ( ) Other: ______________________

2. Gender *

*Mark only one oval.*

- ( ) Male
- ( ) Female

3. Nationality *

*Mark only one oval.*

- ( ) Portuguese
- ( ) Romanian
- ( ) Italian
- ( ) French
- ( ) German
- ( ) Other: ______________________

4. Place of Birth *

*Mark only one oval.*

- Portugal
- Romania
- Italy
- France
- Germany
- Other: ______________________

5. Institution *

*Mark only one oval.*

- University of Porto
- Iuliu Hațieganu University of Medicine and Pharmacy of Cluj-Napoca
- Art and Design University of Cluj-Napoca
- Technical University of Cluj-Napoca
- University Babeș-Bolyai of Cluj-Napoca
- Other: ______________________

6. Course of Studies *

*Mark only one oval.*

- Dentistry
- Art and/or Design
- Engineering
- Other: ______________________

7. Academic Degree *

*Mark only one oval.*

- Student of the last year of Integrated Master
- Student of the 1st year of Integrated Master
- Other: ____________

8. What would motivate you to do an Orthodontic Treatment? Fill in the option that better suits your opinion from 5 to 1, being 5 the maximum (most certainly would) and 1 the minimum (most certainly wouldn't). *

*Mark only one oval per row.*

| | 1 | 2 | 3 | 4 | 5 |
|---|---|---|---|---|---|
| Doctor's advice | | | | | |
| Aesthetic reason | | | | | |
| Functional reason | | | | | |
| Fashion (i.e. my classmates are doing; my siblings did, so I'll do it, etc) | | | | | |

#### 1. IF THIS WAS YOUR MOUTH, WOULD YOU FIND ORTHODONTIC TREATMENT?

For each image, answer as if it were your mouth.
Fill in the option that better suits your opinion from 5 to 1, being 5 the maximum (most certainly would) and 1 the minimum (most certainly wouldn't).

9. Aesthetic component from IOTN 1 *

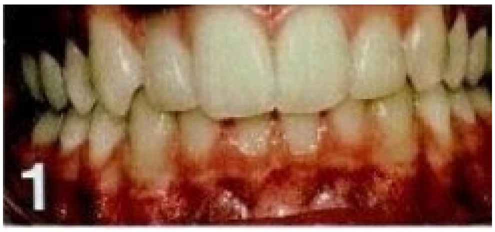


*Mark only one oval per row.*

| | 1 | 2 | 3 | 4 | 5 |
|---|---|---|---|---|---|
| Necessity level | | | | | |

10. Aesthetic component from IOTN 2 *

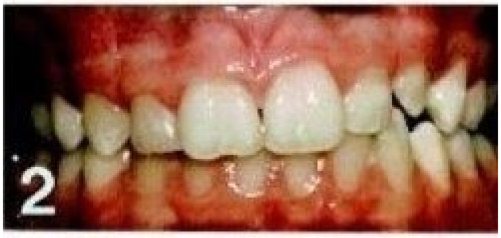


*Mark only one oval per row.*

| | 1 | 2 | 3 | 4 | 5 |
|---|---|---|---|---|---|
| Necessity level | | | | | |

11. Aesthetic component from IOTN 3 *

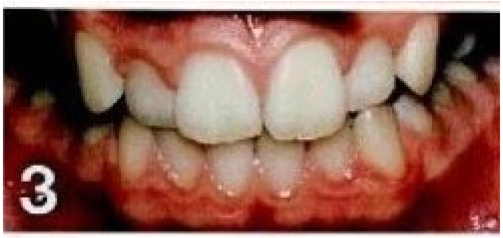


*Mark only one oval per row.*

| | 1 | 2 | 3 | 4 | 5 |
|---|---|---|---|---|---|
| Necessity level | | | | | |

12. Aesthetic component from IOTN 4 *

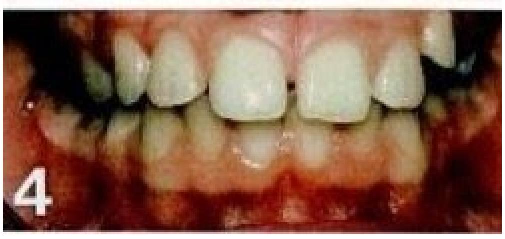


*Mark only one oval per row.*

| | 1 | 2 | 3 | 4 | 5 |
|---|---|---|---|---|---|
| Necessity level | | | | | |

13. Aesthetic component from IOTN 5 *

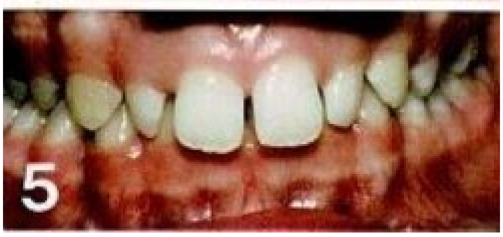


*Mark only one oval per row.*

| | 1 | 2 | 3 | 4 | 5 |
|---|---|---|---|---|---|
| Necessity level | ◯ | ◯ | ◯ | ◯ | ◯ |

14. Aesthetic component from IOTN 6 *

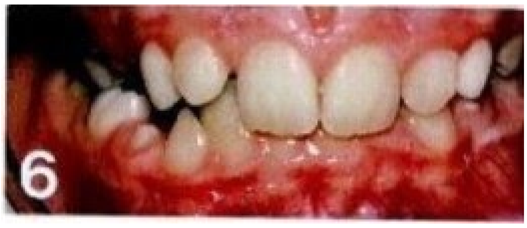


*Mark only one oval per row.*

| | 1 | 2 | 3 | 4 | 5 |
|---|---|---|---|---|---|
| Necessity level | ◯ | ◯ | ◯ | ◯ | ◯ |

15. Aesthetic component from IOTN 7 *

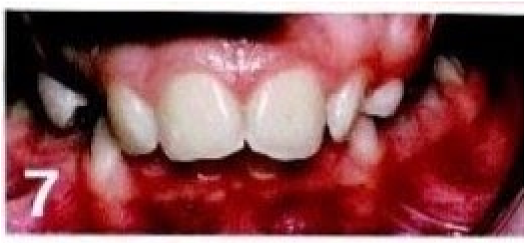


*Mark only one oval per row.*

| | 1 | 2 | 3 | 4 | 5 |
|---|---|---|---|---|---|
| Necessity level | ◯ | ◯ | ◯ | ◯ | ◯ |

16. Aesthetic component from IOTN 8 *

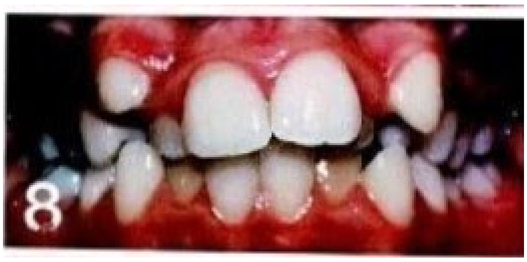


*Mark only one oval per row.*

| | 1 | 2 | 3 | 4 | 5 |
|---|---|---|---|---|---|
| Necessity level | | | | | |

17. Aesthetic component from IOTN 9 *

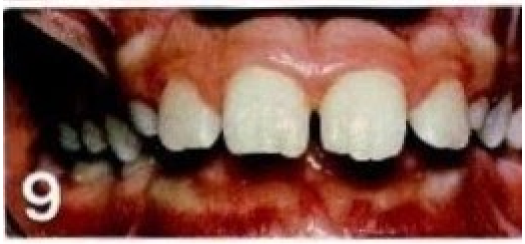


*Mark only one oval per row.*

| | 1 | 2 | 3 | 4 | 5 |
|---|---|---|---|---|---|
| Necessity level | | | | | |

18. Aesthetic component from IOTN 10 *

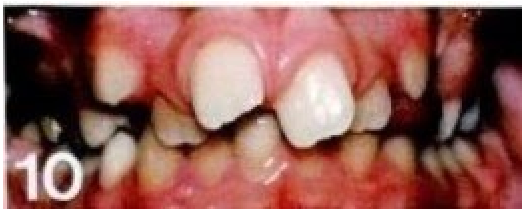


*Mark only one oval per row.*

| | 1 | 2 | 3 | 4 | 5 |
|---|---|---|---|---|---|
| Necessity level | | | | | |

#### 2. IF THIS WAS SOMEONE'S MOUTH, WHO ASKED FOR YOUR OPINION, WOULD YOU RECOMMEND ORTHODONTIC TREATMENT?

For each image, answer as if it were someone else's mouth.
Fill in the option that better suits your opinion from 5 to 1, being 5 the maximum (most certainly needs treatment) and 1 the minimum (most certainly doesn't need treatment).

19. Aesthetic component from IOTN 1 *

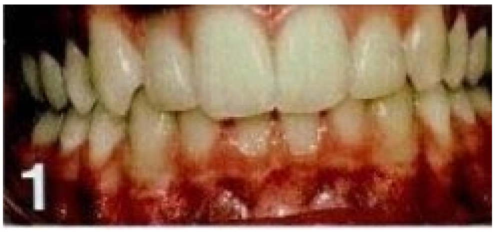


*Mark only one oval per row.*

| | 1 | 2 | 3 | 4 | 5 |
|---|---|---|---|---|---|
| Necessity level | ◯ | ◯ | ◯ | ◯ | ◯ |

20. Aesthetic component from IOTN 2 *

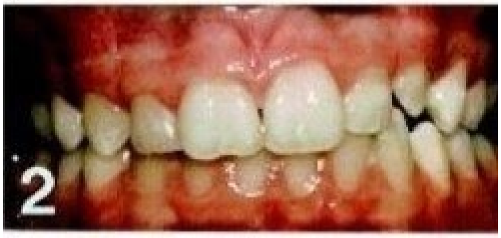


*Mark only one oval per row.*

| | 1 | 2 | 3 | 4 | 5 |
|---|---|---|---|---|---|
| Necessity level | ◯ | ◯ | ◯ | ◯ | ◯ |

21. Aesthetic component from IOTN 3 *

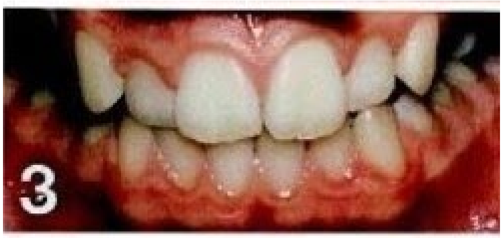


*Mark only one oval per row.*

| | 1 | 2 | 3 | 4 | 5 |
|---|---|---|---|---|---|
| Necessity level | ◯ | ◯ | ◯ | ◯ | ◯ |

22. Aesthetic component from IOTN 4 *

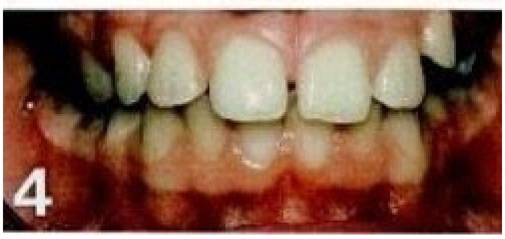


*Mark only one oval per row.*

| | 1 | 2 | 3 | 4 | 5 |
|---|---|---|---|---|---|
| Necessity level | ◯ | ◯ | ◯ | ◯ | ◯ |

23. Aesthetic component from IOTN 5 *

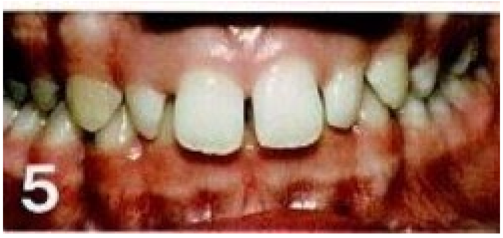


*Mark only one oval per row.*

| | 1 | 2 | 3 | 4 | 5 |
|---|---|---|---|---|---|
| Necessity level | ◯ | ◯ | ◯ | ◯ | ◯ |

24. Aesthetic component from IOTN 6 *

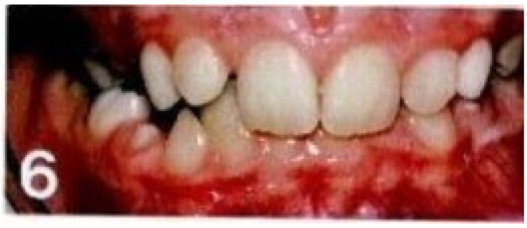


*Mark only one oval per row.*

| | 1 | 2 | 3 | 4 | 5 |
|---|---|---|---|---|---|
| Necessity level | | | | | |

25. Aesthetic component from IOTN 7 *

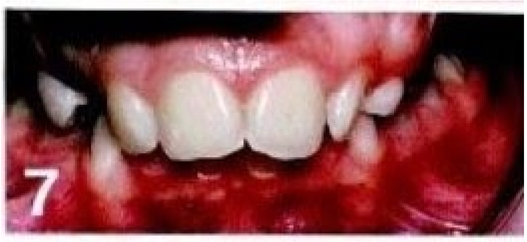


*Mark only one oval per row.*

| | 1 | 2 | 3 | 4 | 5 |
|---|---|---|---|---|---|
| Necessity level | | | | | |

26. Aesthetic component from IOTN 8 *

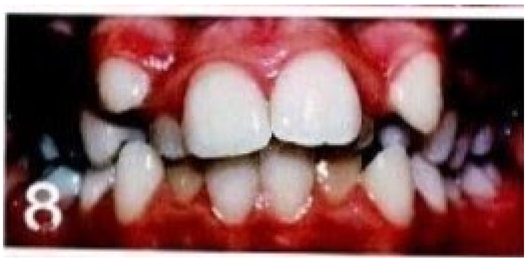


*Mark only one oval per row.*

| | 1 | 2 | 3 | 4 | 5 |
|---|---|---|---|---|---|
| Necessity level | | | | | |

27. Aesthetic component from IOTN 9 *

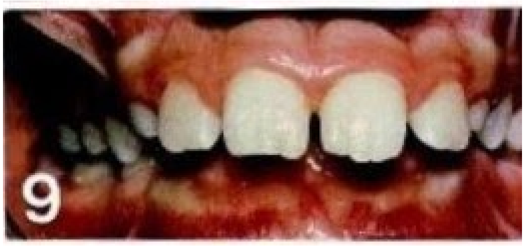


*Mark only one oval per row.*

| | 1 | 2 | 3 | 4 | 5 |
|---|---|---|---|---|---|
| Necessity level | | | | | |

28. Aesthetic component from IOTN 10 *

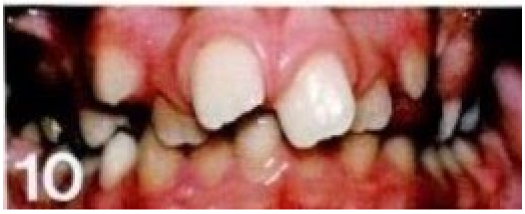


*Mark only one oval per row.*

| | 1 | 2 | 3 | 4 | 5 |
|---|---|---|---|---|---|
| Necessity level | | | | | |

This content is neither created nor endorsed by Google.

Google Forms

Annex 2

Homogeny of the samples for the first part of the study

#### Annex 2

##### Academic Degree * Course of Studies Crosstabulation

| Nationality | | | Course of Studies: Art and/or Design | Dentistry | Ecology and Environment | Engineering | Environmental Engineering | Environmental Sciences | Life Sciences | Medicine | Veterinary Medicine | Total |
|---|---|---|---|---|---|---|---|---|---|---|---|---|
| Portuguese | Academic Degree | Initiated Studies | 2 | 10 | 0 | 3 | | 1 | | 1 | 2 | 19 |
| | | Advanced Studies | 0 | 25 | 1 | 5 | | 0 | | 0 | 0 | 31 |
| | Total | | 2 | 35 | 1 | 8 | | 1 | | 1 | 2 | 50 |
| Romanian | Academic Degree | Initiated Studies | 2 | 10 | | 3 | 1 | | 0 | 1 | 2 | 19 |
| | | Advanced Studies | 0 | 25 | | 5 | 0 | | 1 | 0 | 0 | 31 |
| | Total | | 2 | 35 | | 8 | 1 | | 1 | 1 | 2 | 50 |

Annex 3

Approval of University of Porto’s Data Protection Office (English translated document – 17/12/2020)

**OPINION A-1/2020**

| | |
|---|---|
| **Study** | Aesthetic evaluation of the need for orthodontic treatment – perception among University students |
| **Author** | Luís Emanuel Gonçalves Gomes Alves |
| **Student ID** | 201505219 |
| **Institution** | Faculdade de Medicina Dentária da Universidade do Porto (FMDUP) |
| **Internal Ref.** | 2019121015005409 |

**Summary**

This study was carried out within the curricular unit "Investigation Monography or Clinical Report", integrated in the study plan of the Master Degree in Dental Medicine of FMDUP, with the purposes of evaluating the perception of university students on the need for orthodontic treatment, and determining if that perception is influenced by factors such as age, gender, cultural patterns, the area of study, the educational institution or the academic degree.

The study was implemented through an online questionnaire, which the students of Dental Medicine, Engineering and Fine Arts at the University of Porto, and the students of the corresponding fields at the universities of *Medicină și Farmacie „Iuliu Hațieganu"*, *Babeș-Bolyai*, *Tehnică* and *Artă și Design* in Cluj-Napoca, Romania, were invited to answer.

The perception on the need for orthodontic treatment was esteemed through the aesthetic evaluation of a set of images illustrating malocclusions, considering both the case where the image corresponded to their own mouth and to the mouth of another person. Additionally, the following data were also collected: year of birth, gender, nationality, place of birth, institution, course and academic degree.

**Conclusions**

Considering the set of information collected through the online questionnaire, we are of the opinion that it may be considered anonymous, having as reference the means (human; technological; temporal; financial; etc.) that can reasonably be used to identify a natural person. In this scenario, the data processing involved in this study can be considered exempt from the material scope of Regulation (EU) 2016/679 of the European Parliament and of the Council of 27 April 2016 (General Data Protection Regulation).

In any case, even if the crossing of the information provided by the participants, with additional elements, held by the Data Controller or by third parties, could eventually allow the identification of some of the participants – therefore leading to the qualification of their data as personal data, within the meaning of article 4/1 of the aforementioned Regulation – the corresponding Data Processing was legitimized by their consent, demonstrated explicitly through the voluntary completion of the questionnaire, after being duly informed of the study outlines.

In this context, there were no obstacles found, from the point of view of the legislation in force with regard to the protection of personal data and the free movement of such data, to carry out the study, as described above.

**The Data Protection Officer of the**
**University of Porto**

Assinado por : **SUSANA RODRIGUES PEREIRA**
Num. de Identificação: BI110940423
Data: 2020.12.17 16:39:22 +0000

**Susana Rodrigues Pereira**
