## Supplementary material for "Aesthetic evaluation of the need for orthodontic treatment – Perception among university students": Transparency Declaration

Subject: Transparency Declaration

Research Scientific Article "Aesthetic evaluation of the need for orthodontic treatment - Perception among university students" to be published by Luís Alves et al.

I, Luís Emanuel Gonçalves Gomes Alves, on behalf of Anca Mesaros, Maria João Ponces and Maria Cristina Figueiredo Pollmann, affirm that this manuscript is an honest, accurate, and transparent account of the study being reported; that no important aspects of the study have been omitted; and that any discrepancies from the study as planned (and, if relevant, registered) have been explained.

Porto, December 7th of 2020

The Corresponding Author

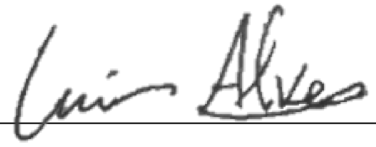

Luís Alves

RUA DR. MANUEL PEREIRA DA SILVA, 4200-392 PORTO - PORTUGAL
www.fmd.up.pt
