## Supplementary material for "Aesthetic evaluation of the need for orthodontic treatment – Perception among university students": Data Sharing Statement

Subject: Data Sharing Statement

Research Scientific Article "Aesthetic evaluation of the need for orthodontic treatment - Perception among university students" to be published by Luís Alves et al.

I, Luís Emanuel Gonçalves Gomes Alves, on behalf of Anca Mesaros, Maria João Ponces and Maria Cristina Figueiredo Pollmann, affirm that the Data on which this manuscript was based is available upon reasonable request.

Porto, December 7th of 2020

The Corresponding Author

Luís Alves

RUA DR. MANUEL PEREIRA DA SILVA, 4200-392 PORTO - PORTUGAL
www.fmd.up.pt
